# Tumor-adjacent B cell infiltration stratifies recurrence risk in localized prostate cancer

**DOI:** 10.64898/2026.08.29.26361718

**Authors:** Binbin Wang, Sumit Mukherjee, Anna Baj, Shana Y. Trostel, Rosina T. Lis, Nichelle C. Whitlock, Anson T. Ku, Kayla E. Heyward, Sumeyra Kartal, Kun Wang, Olga S. Voznesensky, Carla Calagua, Javed Siddiqui, Rabia S. Martin, Lori A. Kollath, Julian Custer, Patrick D. Michael, L. Priya Kunju, Ross Lake, Curtis C. Harris, Kenneth D. Aldape, Lawrence D. True, Curtis Tatsuoka, Elana J. Fertig, Arul Chinnaiyan, Sandeep Gurram, Peter A. Pinto, Adam B. Weiner, Colm Morrissey, Simpa S. Salami, David J. Einstein, Steven P. Balk, Adam G. Sowalsky, Eytan Ruppin

**Affiliations:** Cancer Data Science Laboratory, National Cancer Institute, Bethesda, MD, USA; Epidemiology and Public Health, University of Maryland School of Medicine, Baltimore, MD, USA; Institute for Genome Sciences, University of Maryland School of Medicine, Baltimore, MD, USA; Genitourinary Malignancies Branch, National Cancer Institute, Bethesda, MD, USA; Department of Medicine, Beth Israel Deaconess Medical Center, Boston, MA, USA; Department of Pathology, University of Michigan, Ann Arbor, MI, USA; Department of Urology, University of Washington, Seattle, WA, USA; Urologic Oncology Branch, National Cancer Institute, Bethesda, MD, USA; Laboratory of Cancer Biology and Genetics, National Cancer Institute, Bethesda, MD, USA; Laboratory of Human Carcinogenesis, National Cancer Institute, Bethesda, MD, USA; Laboratory of Pathology, National Cancer Institute, Bethesda, MD, USA; Department of Laboratory Medicine and Pathology, University of Washington, Seattle, WA, USA; Department of Urology, Cedars-Sinai, Los Angeles, CA, USA; Department of Urology, University of Michigan, Ann Arbor, MI, USA

## Abstract

**Background:** Biochemical recurrence (BCR) occurs in 20–40% of men after radical prostatectomy. Existing postoperative recurrence risk tools based on PSA and pathology are clinically useful but show only moderate and variable discrimination, highlighting the need for biomarkers that improve risk stratification and consequent treatment decisions. We hypothesized that the prostate microenvironment, including both the tumor and non-cancerous adjacent tissue, may contain prognostic features associated with adverse postoperative PSA outcomes.

**Methods:** We assembled a cohort of matched tumor-adjacent benign and tumor prostate tissue from 243 men across three institutions to establish a discovery cohort (n=123; 43 postoperative PSA events, 35%) and validation cohort (n=120; 46 events, 38%). For primary binary analyses, a postoperative PSA event included BCR, defined as two consecutive postoperative PSA values ≥0.2 ng/mL, or PSA persistence. We performed RNA sequencing of matched tumor-adjacent benign and tumor tissues, quantified immune signatures, and developed an integrated model combining the adjacent-tissue B-cell signature, preoperative PSA, and radical prostatectomy Gleason score (“BRIGADE”). CAPRA-S-adjusted Cox analyses excluding recurrence-time-0 cases evaluated time to BCR, and CD19 multiplex immunofluorescence provided tissue-level confirmation (n=10).

**Results:** In prostatectomy specimens, tumors from patients without a postoperative PSA event were enriched for B-cell transcriptional programs, whereas tumors from event-positive patients showed elevated proliferation signatures. B-cell-related transcriptional programs were correlated between tumor and adjacent tissue. Tumor-adjacent benign B-cell scores were higher in no-event cases and discriminated postoperative PSA-event status in PCBN discovery (AUC 0.63) and BM validation (AUC 0.81) cohorts, outperforming numerous other immune-related signatures. In CAPRA-S-adjusted Cox sensitivity analyses excluding recurrence-time-0 cases, higher adjacent-tissue B-cell activity was associated with reduced recurrence risk in PCBN (HR 0.42, 95% CI 0.19–0.94; BH-adjusted p=0.035) and BM (HR 0.54, 95% CI 0.30–0.95; BH-adjusted p=0.034). Tissue-based validation showed that CD19⁺ B-cell density in adjacent benign tissue was higher in no-event than event-positive patients (median 0.1145 vs 0.0471; p=0.008). BRIGADE achieved an AUC of 0.68 in cross-validation and 0.83 in independent validation, compared to AUCs of 0.54–0.63 and 0.44–0.78 for the tested clinical predictors, respectively. At the fixed classification threshold, the validation-cohort odds ratio for BRIGADE was 2.75. The adjacent B-cell score remained associated with lower odds of a postoperative PSA event after adjustment for PSA and Gleason score.

**Conclusions:** B-cell infiltration in tumor-adjacent benign prostate tissue may complement existing clinicopathologic models for stratifying adverse postoperative PSA outcomes and subsequent BCR after radical prostatectomy. The transcriptomic signal was recapitulated by CD19-based tissue staining, supporting further development of a pathology-based assay.

## INTRODUCTION

Prostate cancer (PCa) is among the most commonly diagnosed malignancies in men and is a leading cause of global cancer mortality [1]. For patients with localized disease, radical prostatectomy can be curative, but adverse postoperative PSA outcomes include both PSA persistence and later biochemical recurrence (BCR), reflected by a subsequent rise in prostate-specific antigen (PSA). These outcomes occur in a substantial subset of patients and can precede eventual clinical progression [2]. Identifying patients at highest risk of these outcomes is central to postoperative risk stratification and to selecting patients for treatment intensification or de-intensification strategies.

Post-prostatectomy risk prediction remains largely tied to conventional clinicopathologic features such as PSA, ISUP grade group, pathologic stage, margin status, seminal vesicle invasion, and lymph node involvement [3]. These factors contribute to preoperative systems such as D’Amico and CAPRA and to postoperative models such as CAPRA-S and other contemporary nomograms [4–7]. While clinically valuable, external validation of these tools has shown moderate discrimination for BCR across different patient populations, leaving a sizable fraction of patients in an intermediate risk category lacking an optimal oncologic management plan [6, 8–10]. This persistent gap has driven interest in molecular, imaging, and multimodal AI-enabled biomarkers that complement standard variables to improve risk stratification.

Beyond tumor-intrinsic determinants, the neighboring tumor immune microenvironment (TIME) is increasingly recognized as playing an important role in tumor progression and therapeutic response. In PCa, prognostic associations have been reported for multiple components, including CD8+ T cells, NK cells, suppressive myeloid populations, and macrophages, although the roles these immune subsets play are contextually dependent across cohorts and analytic approaches [11–15]. Compared to T cells and myeloid cells, the role of B cells and humoral immune programs is less understood in localized PCa despite increasing recognition that B cells can influence antitumor immunity through antibody-mediated functions, antigen presentation, and cytokine signaling [16]. Recent work further suggests that the prognostic effect of plasma-cell content in PCa may depend on the surrounding inflammatory cytokine milieu, underscoring the importance of both cellular state and tissue context [17]. Understanding these complex immune dynamics may provide opportunities to develop more accurate prognostic biomarkers, while better defining the TIME’s influence on subsequent relapse may inform immunomodulatory strategies.

Importantly, prognostic biology may extend beyond the tumor itself. Histologically benign tissue adjacent to prostate tumors can harbor molecular alterations consistent with a field effect, and features measured in benign fields including nuclear morphology and transcriptional programs have been linked to BCR [18–20]. Consistent with this broader concept, H&E-based analyses of lymphocyte clustering have suggested that spatial immune organization may carry prognostic information, although such approaches do not distinguish B cells from T cells without additional marker-based validation [21]. In this study, we performed transcriptomic profiling of matched tumor and tumor-adjacent benign tissue from a multi-institutional radical prostatectomy cohort and tested whether immune programs in the adjacent microenvironment provide prognostic information for adverse postoperative PSA outcomes and time to subsequent BCR, both alone and in combination with standard clinical variables. Our findings establish the prognostic importance of the immune microenvironment in adjacent benign tissue and provide a foundation for developing immune-based biomarkers to guide personalized treatment decisions in PCa.

## RESULTS

### Clinical predictors and risk scores show variable discrimination across cohorts

We hypothesized that the prostate microenvironment, involving both the tumor and adjacent non-cancerous compartments, may contain features associated with adverse postoperative PSA outcomes. To study this in depth, we assembled a multi-institutional cohort (**Figure 1A**) of 243 patients (**Supplementary Table 1**) treated with radical prostatectomy and collected matched tumor-adjacent benign and tumor tissue for RNA sequencing (see Methods). Patients were drawn from three cohorts: Prostate Cancer Biorepository Network (PCBN; n=123), Beth Israel Deaconess Medical Center (BIDMC; n=84), and University of Michigan (UM; n=36).

**Figure 1.**
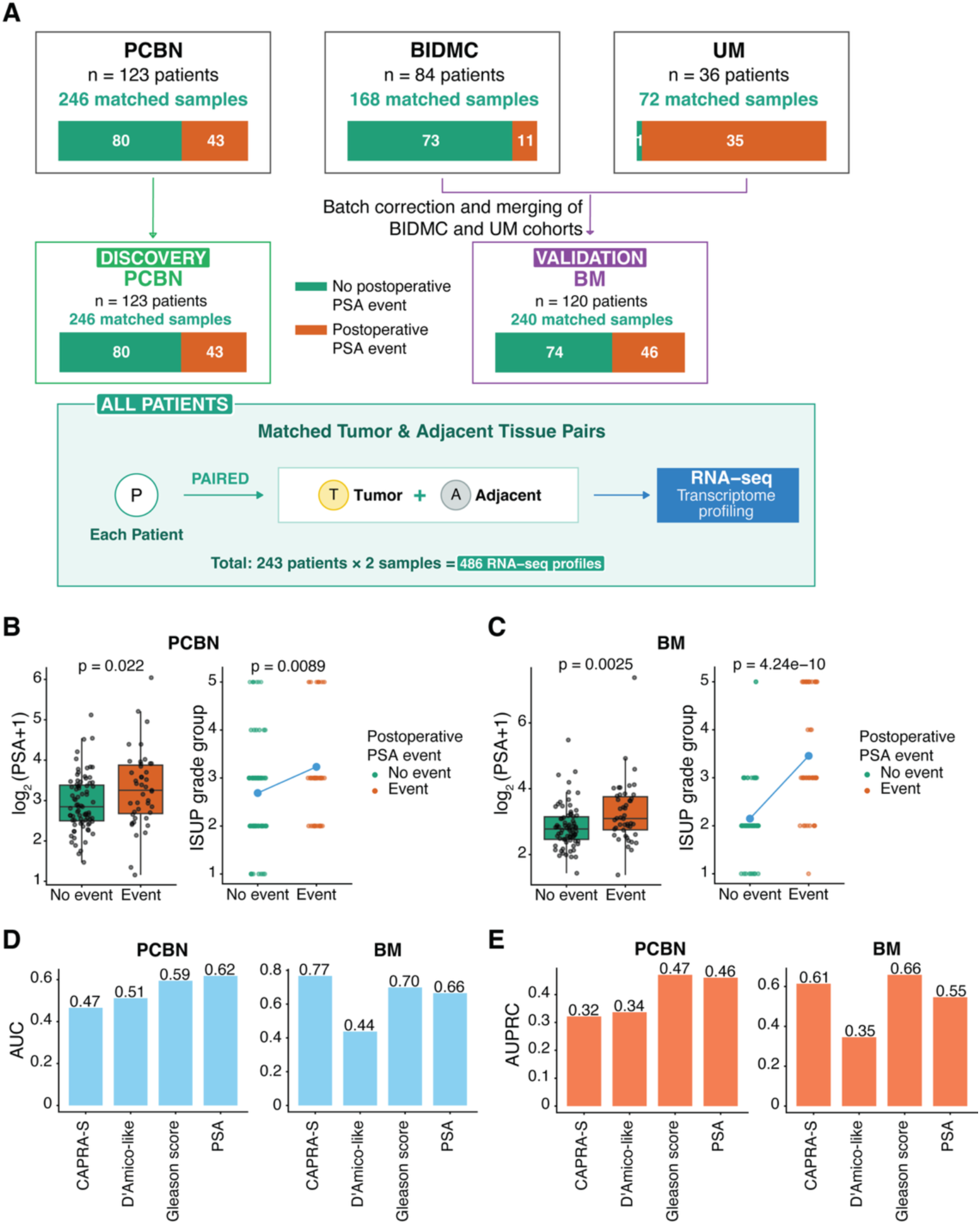
Cohort structure and clinicopathologic predictor performance for postoperative PSA events. **A.** Overview of patient cohorts from three institutions (PCBN, n=123; BIDMC, n=84; UM, n=36). PCBN served as the discovery cohort, and BIDMC and UM transcriptomic data were batch-corrected before merging to form the BM validation cohort. Numbers of patients with and without a postoperative PSA event are indicated for each cohort. For the primary binary analyses, postoperative PSA events included biochemical recurrence or PSA persistence. Matched tumor and tumor-adjacent benign tissue samples were collected from each patient, yielding 486 RNA-seq profiles. **B–C.** Preoperative PSA levels (left) and ISUP grade groups (right) stratified by postoperative PSA-event status in the PCBN (**B**) and BM (**C**) cohorts. For PSA, the center line denotes the median, the box denotes the interquartile range, and whiskers extend to 1.5× the interquartile range. For ISUP grade group, individual patients are shown as points; blue points indicate group means, and the line depicts the trend. P values were determined using a two-tailed Wilcoxon rank-sum test for PSA or the Cochran-Armitage test for trend for ISUP grade group. **D.** Area under the receiver operating characteristic curve (AUC) for postoperative PSA-event classification using PSA, Gleason score, CAPRA-S, and the postoperative D’Amico-like risk classification in PCBN and BM cohorts. **E.** Area under the precision-recall curve (AUPRC) for the same predictors in PCBN and BM cohorts. Preoperative PSA was available for 122 of 123 PCBN patients and all 120 BM patients.

For the primary binary analyses, the outcome was termed a postoperative PSA event. Biochemical recurrence was defined as two consecutive postoperative PSA measurements ≥0.2 ng/mL. Within PCBN, 15 event-positive cases were annotated as persistent disease or had recurrence time recorded as 0 and were therefore considered PSA persistence rather than biochemical recurrence. These cases were retained in the primary binary analyses but excluded from analyses requiring an evaluable interval to recurrence.

BIDMC and UM differed markedly in outcome distribution and clinicopathologic risk profile. BIDMC was enriched for no-event cases (73/84, 86.9%) and lower-grade disease, whereas UM was enriched for event-positive cases (35/36, 97.2%), higher preoperative PSA, higher ISUP grade group, and more advanced pathologic stage (**Supplementary Table 1**). To obtain an independent validation cohort with representation of both outcomes, we combined BIDMC and UM into BM (n=120; 74 no event, 46 event), while PCBN served as the discovery cohort (n=123; 80 no event, 43 event). Amongst patients included in the time-to-event analyses, median follow-up for those without BCR was 50.7 months (range, 3.5–148.3 months), whereas interval BCR occurred at a median of 21.7 months (range, 1.9–144.9 months) after surgery.

As a benchmark, we evaluated PSA and Gleason score, reported here also as ISUP grade group, by postoperative PSA-event status and assessed discrimination using individual variables (PSA and ISUP grade group) and composite classifications (postoperative D’Amico-like and CAPRA-S). Across both PCBN and BM, event-positive cases exhibited higher median PSA levels and higher ISUP grade group (**Figure 1B–C**). Given the outcome imbalance within BM, we benchmarked PSA, ISUP grade group, CAPRA-S, and postoperative D’Amico-like risk classification before evaluating transcriptomic predictors (**Figure 1D–E**). Predictive performance of the clinical variables and risk scores varied across cohorts: in PCBN, AUC values ranged from 0.47–0.62, while in BM they ranged from 0.44–0.77, with corresponding AUPRC values of 0.32–0.47 and 0.35–0.66, respectively (**Figure 1D–E**). This variability prompted our evaluation of molecular features in tumor and adjacent tissue to identify biomarkers with more consistent performance across distinct cohorts.

### Tumor-adjacent benign transcriptional analyses implicate B cell programs

Matched tumor-adjacent benign and tumor tissue samples were collected from all patients during prostatectomy and subjected to RNA sequencing. For cross-cohort analyses, ComBat-seq was applied separately to the benign and tumor matrices to reduce cohort-associated technical variation (**Supplementary Figure 1**). Using the original, uncorrected PCBN count data for discovery, we compared adjacent benign transcriptomes between patients with and without a postoperative PSA event. At the prespecified thresholds of fold-change >1.5 and FDR <0.1, 20 transcripts were differentially expressed, of which 10 of the 15 transcripts with higher expression in the no-event group were related to immunoglobulin and B-cell biology (**Figure 2A**). We therefore curated a set of 17 published immune-related signatures spanning immune-cell, antigen-presentation, cytolytic, stromal, and immune-regulatory programs and tested whether their ssGSEA enrichment scores could distinguish postoperative PSA-event status. Among these signatures, the B-cell gene expression signature reported by Budczies et al. [22] showed the strongest discrimination in tumor-adjacent benign transcriptomes in both the PCBN and BM cohorts (**Figure 2B**; **Supplementary Table 2**). We refer to this ssGSEA enrichment score in adjacent benign tissue as the adjacent benign B-cell signature score.

**Figure 2.**
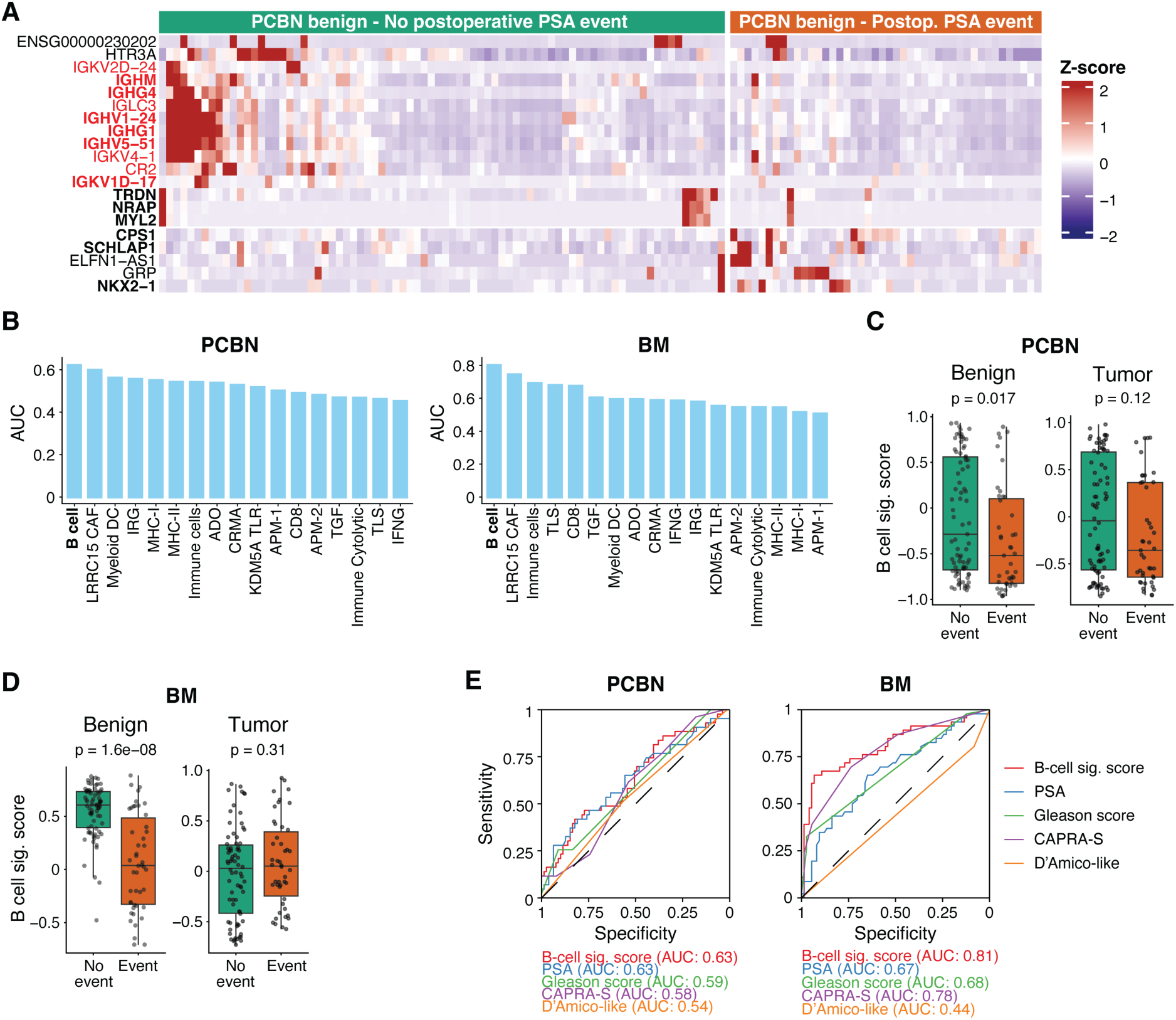
Transcriptomic programs in tumor-adjacent benign tissue. **A.** Heatmap of differentially expressed genes (fold-change > 1.5, FDR < 0.1) comparing tumor-adjacent benign prostatectomy tissues from patients with and without a postoperative PSA event in the PCBN cohort (n=123). Genes shown in red are associated with B-cell biology, and genes in boldface also passed the more stringent FDR threshold of q < 0.05. **B.** Area under the receiver operating characteristic curve (AUC) for a curated set of immune-related gene signatures applied to tumor-adjacent benign tissues from the PCBN and BM cohorts. PubMed IDs and available gene members are provided in **Supplementary Table 2**. **C–D.** B-cell signature enrichment scores in tumor-adjacent benign and tumor tissues from the PCBN (**C**) and BM (**D**) cohorts, stratified by postoperative PSA-event status. P values were determined using two-tailed Wilcoxon rank-sum tests. Box plots indicate the median (center line), interquartile range (box), and whiskers extending to 1.5× the interquartile range. **E.** ROC curves for the adjacent benign B-cell signature score, PSA, Gleason score, CAPRA-S, and the postoperative D’Amico-like risk classification in the PCBN and BM cohorts.

To determine whether this immune signal was also present in tumor tissue, we next compared tumor transcriptomes by postoperative PSA-event status. Tumors from no-event patients similarly displayed enrichment for immune biology and B-cell programs, whereas tumors from event-positive patients were enriched for proliferation-associated programs (**Supplementary Figure 2A–B**). Immune-related transcripts, including B-cell-related genes, were strongly correlated between matched tumor and tumor-adjacent benign tissues (**Supplementary Figure 2C–E**). The adjacent benign and tumor B-cell signature scores were positively correlated across matched samples in PCBN (Pearson r=0.42, p=1.1×10⁻⁶) and BM (r=0.24, p=0.0072; **Supplementary Figure 2F**), although only the adjacent benign score discriminated postoperative PSA-event status; the tumor B-cell signature score did not significantly differ by postoperative PSA-event status in either cohort (**Figure 2C–D**). By contrast, the adjacent benign B-cell signature score differed between no-event and event-positive cases in both the PCBN discovery cohort (p=0.017; **Figure 2C**) and the BM validation cohort (p=1.6×10⁻⁸; **Figure 2D**). As a standalone predictor, the adjacent benign B-cell signature score discriminated postoperative PSA-event status with an AUC of 0.63 in PCBN and 0.81 in BM (**Figure 2B,E**). In PCBN, its AUC was similar to that of PSA and higher than those of Gleason score, CAPRA-S, and the postoperative D’Amico-like classification; in BM, its AUC was higher than those of all tested clinical comparators (**Figure 2E**).

Because the PSA-persistence annotations were specific to PCBN, we repeated the PCBN immune-signature analysis after excluding the 15 patients with PSA persistence or recurrence time recorded as 0. In this sensitivity analysis (n=108; 28 biochemical-recurrence events), the B-cell signature remained amongst the top-ranked immune signatures tested. The adjacent benign B-cell signature score remained higher in patients without subsequent BCR (p=0.017), whereas the corresponding tumor score did not differ by subsequent BCR status (p=0.12; **Supplementary Figure 3**). Thus, the adjacent benign B-cell signature emerged as a consistently top-ranked immune feature associated with favorable postoperative PSA outcomes across cohorts.

### Adjacent-tissue B-cell activity remains associated with time to recurrence and broader pathway programs

To evaluate the relationship between immune signatures and time to biochemical recurrence while accounting for established clinical risk, we next performed CAPRA-S-adjusted Cox proportional hazards analyses as a sensitivity analysis. After exclusion of patients with an event or censoring time recorded as 0 and the one PCBN patient with missing preoperative PSA, these models included 107 PCBN patients with 28 biochemical-recurrence events and 118 BM patients with 45 events. Higher adjacent benign B-cell signature scores were associated with a lower hazard of recurrence in both PCBN (HR 0.42, 95% CI 0.19–0.94, BH-adjusted p=0.035; **Figure 3A**) and BM (HR 0.54, 95% CI 0.30–0.95, BH-adjusted p=0.034; **Figure 3B**). In tumor tissue, the B-cell signature was not significantly associated with recurrence in PCBN (HR 0.65, 95% CI 0.34–1.27, BH-adjusted p=0.208; **Figure 3C**) or BM (HR 1.77, 95% CI 0.85–3.70, BH-adjusted p=0.129; **Figure 3D**).

**Figure 3.**
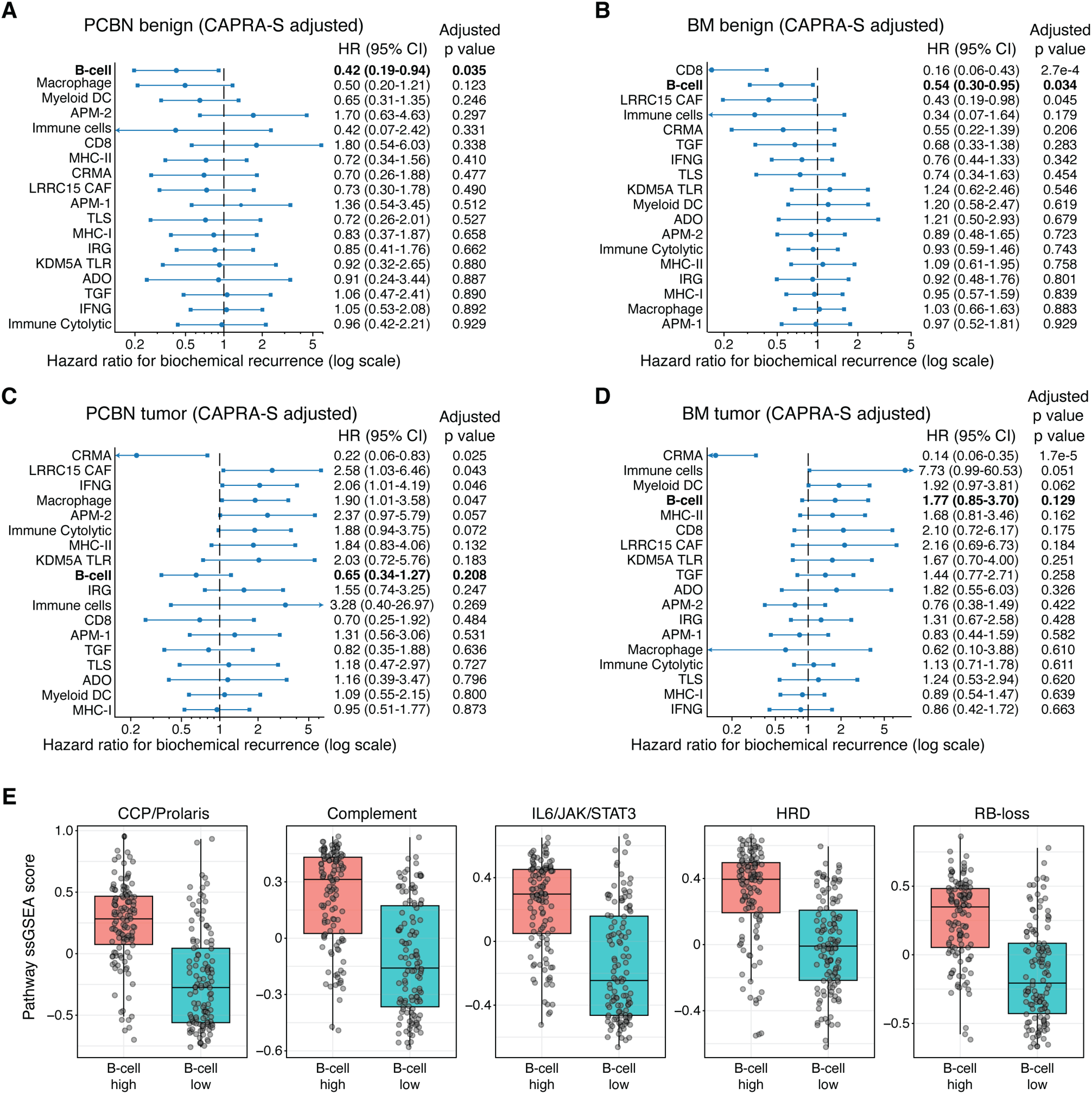
CAPRA-S-adjusted Cox and pathway analyses of B-cell-associated programs. **A–B.** CAPRA-S-adjusted Cox proportional hazards analyses of continuous immune-related ssGSEA signature scores in tumor-adjacent benign tissue from the PCBN (**A**; n=107, 28 biochemical-recurrence events) and BM (**B**; n=118, 45 biochemical-recurrence events) cohorts. The PCBN CAPRA-S-adjusted models excluded one additional patient for whom preoperative PSA was unavailable. **C–D.** Corresponding CAPRA-S-adjusted Cox analyses of immune-related ssGSEA signature scores in tumor tissue from the PCBN (**C**) and BM (**D**) cohorts. Points denote hazard ratios, and horizontal lines denote 95% confidence intervals; estimates are shown on a logarithmic scale. The vertical line indicates a hazard ratio of 1. Arrows indicate confidence intervals extending beyond the displayed axis limits. B-cell signature rows are shown in boldface. P values shown are Wald p values from the CAPRA-S-adjusted Cox models, adjusted across the immune signatures tested using the Benjamini–Hochberg method. **E.** Pathway ssGSEA scores in tumor-adjacent benign tissues from the entire cohort (n=243) stratified into B-cell-high and B-cell-low groups using the median adjacent benign B-cell signature score. Box plots indicate the median, interquartile range, and whiskers extending to 1.5× the interquartile range; individual samples are shown as points. Group comparisons were performed using two-tailed Wilcoxon rank-sum tests. Complete pathway comparison and continuous-correlation results are provided in **Supplementary Tables 3 and 4**.

We further examined the broader pathway programs associated with adjacent benign B-cell activity. Adjacent benign samples from the full cohort (n=243) were stratified into B-cell-high and B-cell-low groups using the median adjacent benign B-cell signature score. B-cell-high adjacent tissues showed higher scores for inflammatory and immune-related pathways, including Complement and IL6/JAK/STAT3 signaling, as well as several signatures associated with aggressive prostate cancer biology, including CCP/Prolaris, HRD, and RB-loss (**Figure 3E**; **Supplementary Figure 4**; **Supplementary Table 3**). Continuous adjacent benign B-cell signature scores were positively correlated with several of the same pathways, including CCP/Prolaris, IL6/JAK/STAT3 signaling, RB-loss, Complement, and HRD signatures (**Supplementary Table 4**). Despite the concurrent enrichment of both immune/inflammatory and aggressive prostate cancer-associated transcriptional programs in B-cell-high adjacent tissues, the favorable prognostic association of adjacent-tissue B-cell activity was maintained after CAPRA-S adjustment, prompting us to test whether combining this signal with standard clinical variables could further improve classification.

### BRIGADE integrates adjacent B cell signatures with PSA and Gleason score

Given the reproducible performance of the adjacent benign B-cell signature across the primary binary analyses and the time-to-event sensitivity analysis, we next asked whether integrating the signature with PSA and Gleason score would improve postoperative PSA-event classification. Using the 122 PCBN patients with complete preoperative PSA data as the training cohort, we fit a regularized logistic regression model with three prespecified predictors: adjacent benign B-cell signature score, preoperative PSA, and Gleason score (**Figure 4A**). This composite score, “**B** cell **R**isk **I**ndex using **G**leason score, **A**djacent-tissue signature, and **D**iagnostic PSA for biochemical recurrenc**E**” (BRIGADE), was then evaluated without refitting in the independent BM validation cohort. BRIGADE achieved a cross-validated AUC of 0.68 and AUPRC of 0.48 in PCBN and an AUC of 0.83 and AUPRC of 0.79 in BM (**Figure 4B–C**).

**Figure 4.**
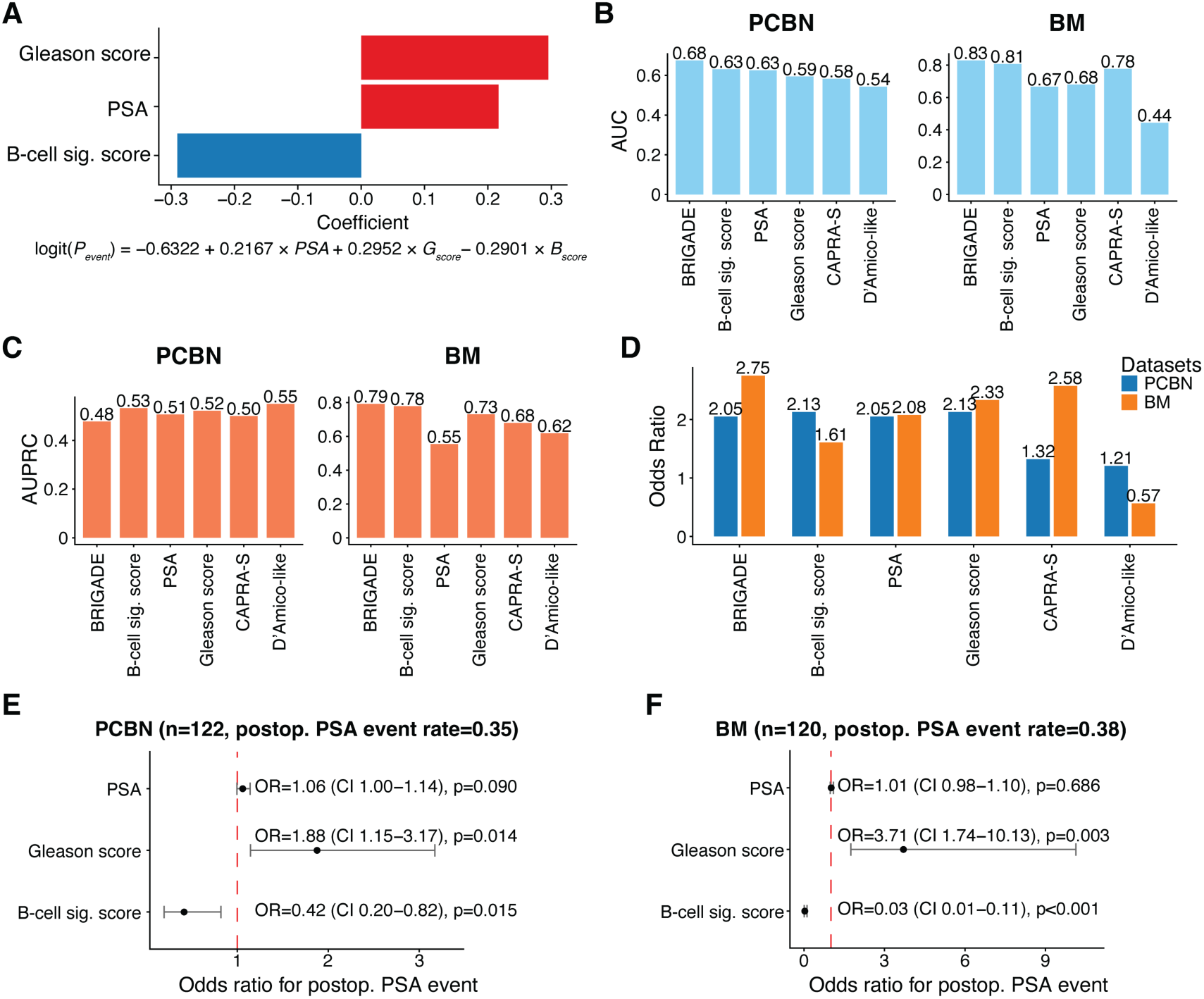
BRIGADE model integrating adjacent-tissue B-cell signature with PSA and Gleason score. **A.** Elastic-net logistic regression coefficients for the three prespecified BRIGADE features: adjacent B-cell signature score, preoperative PSA, and Gleason score. The equation shows the fitted logistic model for the probability of a postoperative PSA event. **B–C.** Areas under the ROC curves (**B**) and precision-recall curves (**C**) for BRIGADE and comparator predictors including the adjacent benign B-cell signature score, PSA, Gleason score, CAPRA-S, and the postoperative D’Amico-like risk classification. Performance is shown for cross-validation in PCBN complete-case cohort (n=122) and independent validation in BM (n=120). **D.** Odds ratios for a postoperative PSA event comparing predictor-defined high-risk and low-risk groups in the PCBN and BM cohorts. Predictor-specific classification thresholds were selected using Youden’s index, and odds ratios and 95% confidence intervals were estimated by logistic regression. **E–F.** Forest plots depicting odds ratios and 95% confidence intervals from multivariable logistic regression models including PSA, Gleason score, and the adjacent benign B-cell signature score in the PCBN (**E**) and BM (**F**) cohorts. The red dashed line indicates an odds ratio of 1.

For threshold-based classification, we selected a fixed BRIGADE threshold (0.361) in the PCBN training cohort using the Youden’s index method, which maximizes sensitivity and specificity (**Supplementary Figure 5A**). We then applied the same cutoff to the independent test cohort, BM. At this threshold, BRIGADE yielded odds ratios of 2.05 in PCBN and 2.75 in BM (**Figure 4D**; **Supplementary Figure 5B–C**). Corresponding threshold-based classifications for the adjacent benign B-cell signature score and the clinical predictors are shown in **Figure 4D** and **Supplementary Figure 5D–E**. Moreover, in logistic regression models adjusting for PSA and Gleason score, the adjacent-tissue B cell signature remained independently associated with lower odds of a postoperative PSA event in both cohorts (PCBN OR 0.42, 95% CI 0.20–0.82, p=0.015; BM OR 0.03, 95% CI 0.01–0.11, p<0.001) (**Figure 4E–F**), with no consistently strong associations between the B cell signature and either PSA or Gleason score (**Supplementary Figure 6A–B**).

Because the transcriptomic findings implicated B-cell biology beyond cellular abundance alone, we also explored adjacent-tissue B-cell receptor repertoire clonality. Patients with higher Gini indices, reflecting greater clonal inequality and lower repertoire diversity, showed a trend toward longer BCR-free survival (log-rank p=0.089; **Supplementary Figure 7A**). Collectively, these results support the transcriptomic measurement of B-cell infiltration as a prognostic marker that provides information beyond PSA and Gleason score.

### CD19 immunofluorescence confirmation of adjacent-tissue B cell enrichment

Finally, to determine whether the transcriptome-inferred B-cell signature reflected B-cell abundance at the tissue level, we performed multiplex CD19 immunofluorescence in a 10-patient verification cohort (5 no-event and 5 event-positive cases). In the no-event cases, CD19⁺ B cells were observed in adjacent tissue as lymphoid aggregates and as scattered individual cells throughout the stromal compartment (**Figure 5A**). By contrast, event-positive samples showed sparse or absent CD19⁺ cells in the adjacent tissue (**Figure 5A**). Similarly, representative tumor regions showed prominent CD19⁺ B-cell infiltration in no-event cases and comparatively sparse staining in event-positive cases (**Figure 5A**).

**Figure 5.**
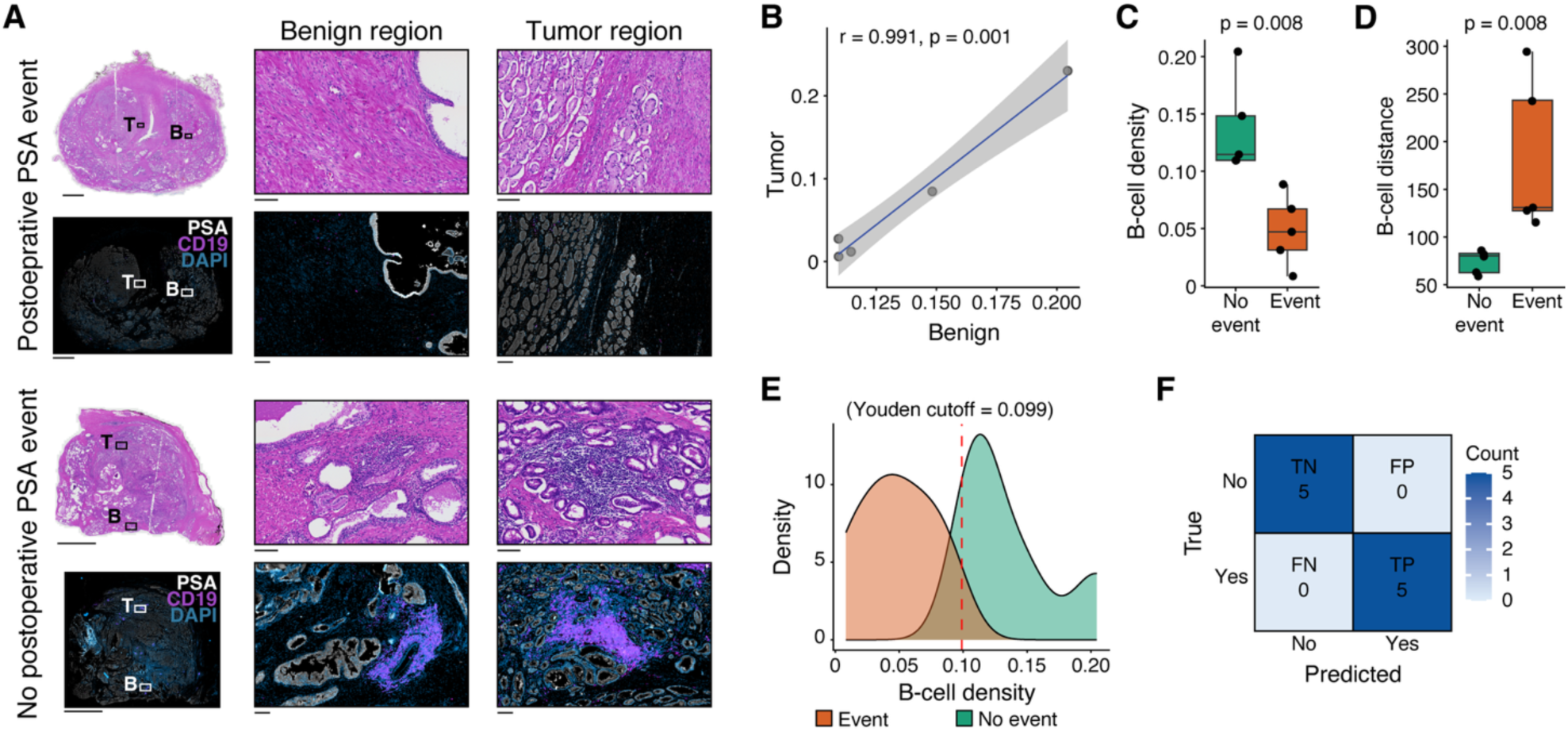
Multiplex immunofluorescence quantification of CD19^+^ B cells in tumor and tumor-adjacent benign tissue. **A.** H&E and multiplex immunofluorescence images from representative cases with (top) or without (bottom) a postoperative PSA event (total n=10; 5 event and 5 no-event cases). For each case, H&E images are shown in the upper row and multiplex immunofluorescence images in the lower row. Whole-slide views are shown at left, with magnified benign and tumor regions shown at center and right, respectively. Tumor (T) and tumor-adjacent benign (B) regions are indicated on the whole-slide views. Multiplex immunofluorescence shows PSA in white, CD19 in magenta, and DAPI in cyan. Scale bars: whole-slide views, 5 mm; magnified fields, 100 μm. **B.** Pearson correlation of normalized CD19⁺ B-cell density between matched tumor and tumor-adjacent benign tissues in cases with matched specimens (n=5). **C.** Normalized CD19⁺ B-cell density in tumor-adjacent benign tissue, stratified by postoperative PSA-event status. **D.** B-cell nearest-neighbor distance stratified by postoperative PSA-event status. For panels C–D, P values were determined using two-tailed Wilcoxon rank-sum tests; box plots indicate the median, interquartile range, and whiskers extending to 1.5× the interquartile range. **E.** Kernel density estimates of normalized CD19⁺ B-cell density in event and no-event cases. The dashed line indicates the classification cutoff of 0.099 selected using Youden’s index. **F.** Confusion matrix for classification of postoperative PSA-event status using the cutoff identified in panel **E**. TP, event-positive patient classified as high risk; FN, event-positive patient classified as low risk; FP, no-event patient classified as high risk; and TN, no-event patient classified as low risk.

We then applied automated image analysis to quantify normalized CD19⁺ B-cell density and nearest-neighbor distance. Normalized CD19⁺ B-cell density was strongly correlated between matched adjacent benign and tumor compartments (Pearson r=0.991, p=0.001; **Figure 5B**). Adjacent benign tissue from no-event patients had higher normalized CD19⁺ B-cell density than tissue from event-positive patients (median 0.1145 versus 0.0471; two-tailed Wilcoxon rank-sum p=0.00794; **Figure 5C**), whereas event-positive cases had greater B-cell nearest-neighbor distance (p=0.00794; **Figure 5D**). Time-to-event visualizations of these same density and nearest-neighbor measurements in the 10-patient cohort are shown in **Supplementary Figure 7B–C**.

We applied a normalized B-cell density cutoff of 0.099, selected using Youden’s index, to classify postoperative PSA-event status (**Figure 5E**). At this cutoff, all five no-event cases and all five event-positive cases were correctly classified in this tissue-verification cohort (**Figure 5F**). Taken together, these immunofluorescence findings confirm the transcriptomic B-cell signal at the tissue level, with greater CD19⁺ B-cell infiltration in adjacent tissue associated with favorable clinical outcomes following radical prostatectomy.

## DISCUSSION

In this multi-institutional study of a uniquely matched cohort of 243 PCa patients treated with radical prostatectomy, we identify B-cell infiltration in tumor-adjacent benign prostate tissue as a robust prognostic biomarker of adverse postoperative PSA outcomes. Across independent cohorts, adjacent-tissue B-cell signatures discriminated postoperative PSA events more consistently than standard clinicopathologic variables and commonly used clinical risk models and remained associated with time to biochemical recurrence after CAPRA-S adjustment among patients with an evaluable interval to recurrence. Integrating the adjacent-tissue B-cell signature with PSA and Gleason score in a simple composite model (BRIGADE) improved performance in an external validation cohort, supporting the hypothesis that clinically relevant prognostic information is encoded not only within the tumor transcriptome but also within the surrounding benign microenvironment.

Despite increasing data on a subset of immunogenic PCa, our findings emphasize that humoral immune programs may carry clinically meaningful prognostic signal in localized disease. B cells can participate in antitumor immunity through antibody-mediated effector functions, antigen presentation, and immune modulation, but the impact of B cells can be contextually dependent upon both cellular state and tissue location [16]. In the present study, we applied bulk RNA-seq and CD19-based staining that detected a B-cell signal but did not further resolve B-cell subsets, plasma-cell differentiation, activation state, or spatial architecture including tertiary lymphoid structures. Reconstruction of B-cell receptor repertoires suggested a trend towards lower diversity among patients with longer BCR-free survival, although this association did not reach statistical significance. Nonetheless, our concordant transcriptomic and tissue-level data imply that the abundance and spatial organization of B cells within tumor-adjacent tissue tracks with favorable outcomes, nominating this immune axis for deeper investigation. This context-dependent interpretation is consistent with prior prostate cancer work showing that the prognostic effect of plasma-cell content may depend upon the surrounding inflammatory cytokine milieu [17], and with similar findings in head and neck squamous cell carcinoma, in which tumor and blood B-cell abundance outperformed several established immunotherapy-response signatures and tracked with a more favorable antitumor microenvironment [23].

Our additional time-to-event and pathway analyses provide important biological context for the adjacent-tissue B-cell finding. The primary classification analysis included patients with PSA persistence, whereas the time-to-event analysis tested whether the adjacent B-cell signal also preceded subsequent BCR among patients with an evaluable interval to recurrence. The persistence of a statistically significant B-cell association in the latter analysis indicates that the association is not driven solely by the inclusion of immediate postoperative failure. Notably, B-cell-high adjacent tissue was associated not only with immune and inflammatory pathways, including Complement and IL6/JAK/STAT3 signaling, but also with transcriptional programs linked to aggressive prostate cancer biology, including CCP/Prolaris, RB-loss, and HRD signatures. This juxtaposition suggests that the favorable association of adjacent-tissue B-cell activity reflects an active host antitumor response rather than simply identifying patients with less aggressive disease. These findings support a model in which a B-cell-rich adjacent immune field reflects stronger host antitumor capability; tumors that progress in B-cell-high patients may require more aggressive molecular adaptations to overcome local immune pressure, whereas tumors in B-cell-low patients may progress in the setting of weaker adjacent-tissue immune surveillance.

Notably, our observation that adjacent benign tissue performed as well as, or better than, tumor tissue aligns with broader literature suggesting that tissue outside the malignant epithelium can harbor clinically informative biology. Field cancerization has been described in the prostate amongst other solid tumor types, with molecular alterations detectable in morphologically benign regions that surround tumors [18, 24]. In colorectal cancer, transcriptomes from tumor-adjacent normal tissue were more informative than tumors for predicting recurrence, supporting the idea that histologically nonmalignant tissue can encode prognostic signal [25]. Indeed, benign glands including nuclear architecture and gene expression profiles can predict biochemical recurrence following prostatectomy [19, 20], and spatial and longitudinal patterns of CD20^+^ and CD3^+^ lymphocytes across benign biopsy, tumor-adjacent benign glands and malignant glands have also been associated with biochemical recurrence, indicating that immune changes in benign compartments are not entirely unexpected [26]. More broadly, recent prostate studies have identified other microenvironment-derived recurrence signals, including a recurrence-associated endothelial signature and collagen-based molecular models that improve prediction of biochemical recurrence after radical prostatectomy [27, 28]. Our current results extend this emerging framework by specifically implicating a B-cell transcriptional program in histologically benign tumor-adjacent prostate tissue and by linking that signal to an orthogonal CD19-based tissue phenotype and a time-to-recurrence association.

Improved postoperative risk stratification may ultimately help identify patients for whom early salvage radiotherapy or the addition of androgen-deprivation therapy to salvage radiotherapy should be considered, while allowing others to remain under close monitoring and avoiding overinterpretation of biomarkers that have not yet been prospectively validated [29–31]. BRIGADE was intentionally designed to integrate a single adjacent-tissue immune signature with variables already collected in routine care, including PSA and Gleason score. While RNA-based approaches could support implementation, our histologic analysis suggests that B-cell quantification may provide a practical pathologic readout of the adjacent-tissue signal. Because CD20 and CD79A are more commonly used in diagnostic pathology than CD19, future studies should determine whether these routinely available markers reproduce the CD19-based association. Further work should prioritize standardized sampling of tumor-adjacent benign tissue, robust scoring approaches, and evaluation of whether adjacent-tissue B-cell density adds value to established post-prostatectomy decision tools, including commercially available molecular classifiers [32].

Our study has several limitations. First, bulk transcriptomic profiling and CD19-based immunofluorescence cannot fully distinguish B-cell subsets, plasma-cell differentiation states, antigen specificity, TLS organization, and B-cell receptor clonality. Second, all three cohorts were retrospective and differed in recurrence incidence and follow-up. In particular, the independent BM validation cohort was created by merging BIDMC and UM, which had highly divergent recurrence frequencies and clinicopathologic risk profiles. Third, our primary endpoint combined biochemical recurrence and PSA persistence, allowing clinically aggressive persistent cases to contribute to biomarker discovery. Although our Cox models excluded cases with PSA persistence, these were supportive sensitivity analyses. Finally, although BCR is clinically meaningful and can precede disease progression, it is not a validated surrogate for metastasis or cancer-specific mortality; longer follow-up and metastasis-specific endpoints will therefore be required to establish the clinical utility of this biomarker.

Overall, our findings position B-cell infiltration into the tumor-adjacent benign compartment of localized PCa as a prognostic biomarker of adverse postoperative PSA outcomes and subsequent biochemical recurrence after radical prostatectomy. Our data support the view that tumor-adjacent benign tissue is clinically informative and identify a B-cell program within that compartment as a tractable signal for further study. The association of adjacent-tissue B-cell activity with favorable outcomes despite enrichment of aggressive prostate cancer-associated transcriptional programs suggests that this biomarker may capture host immune capability beyond tumor-intrinsic risk alone. Clarifying potential interactions between adjacent-tissue immunity, tumor genomics, and early drivers of metastatic spread may further improve our mechanistic understanding of this novel biomarker. Future studies should assess generalizability across treatment settings and determine whether adjacent-tissue immune biomarkers add value to existing nomograms and commercial genomic classifiers for risk-adapted treatment intensification or de-intensification.

## METHODS

### Patient cohorts and sample collection

Transcriptomic analyses included three institutional cohorts of patients treated by radical prostatectomy: the Prostate Cancer Biorepository Network (PCBN; Seattle, WA), Beth Israel Deaconess Medical Center (BIDMC; Boston, MA), and the University of Michigan (UM; Ann Arbor, MI). An additional National Cancer Institute (NCI; Bethesda, MD) cohort provided tissue for the multiplex immunofluorescence verification analysis. All patients provided informed consent, and the study was approved by institutional review boards at each participating institution.

For the PCBN, BIDMC and UM cohorts, clinical data were collected including age at diagnosis, preoperative PSA level, Gleason score, pathologic tumor stage, postoperative PSA-event status, and follow-up information. For the primary binary analyses, a postoperative PSA event was defined as either biochemical recurrence or PSA persistence. Biochemical recurrence was defined as two consecutive postoperative PSA measurements ≥0.2 ng/mL following radical prostatectomy. Within PCBN, 15 event-positive patients annotated as having persistent disease (postoperative PSA not decreasing to <0.1 ng/mL) or with time to event recorded as 0 were classified as having PSA persistence and were retained as event-positive in the primary binary analyses. For time-to-event analyses, patients with an event or censoring time recorded as 0 were excluded. Preoperative PSA was unavailable for one PCBN patient. No missing clinical values were imputed. This patient was retained in transcriptomic and other analyses that did not require preoperative PSA but was excluded from analyses incorporating preoperative PSA or PSA-derived clinical scores, including the postoperative D’Amico-like risk classification, CAPRA-S, BRIGADE, and PSA-adjusted multivariable models. Time was measured from radical prostatectomy to biochemical recurrence or last clinical follow-up.

Matched tumor and adjacent tissue samples were collected from all patients during radical prostatectomy. Frozen prostatectomy specimens from PCBN were embedded in OCT, and fixed prostatectomy specimens from BIDMC and UM were embedded in paraffin. Specimens were sectioned onto glass slides and stained with H&E or shipped as ribbon curls in microfuge tubes. After review by a genitourinary pathologist to confirm the presence of tumor cells, ribbon curls were processed using the RNeasy Plus Mini Kit (Qiagen) or the RNeasy FFPE Mini Kit (Qiagen) according to the manufacturer’s instructions to extract total RNA. All tissue samples were reviewed by board-certified genitourinary pathologists to confirm diagnosis and tissue classification.

### RNA sequencing and data processing

RNA was assembled into strand-specific, paired-end, Illumina-compatible sequencing libraries using the NEBNext Ultra II Directional RNA Library Prep Kit (New England Biolabs) with the NEBNext rRNA Depletion Kit (New England Biolabs). Libraries were sequenced on Illumina NovaSeq 6000 S4 and NovaSeq X Plus 25B flowcells to generate 2 × 100-150 bp reads with a target depth of at least 40 million read pairs per sample.

Raw sequencing reads in FASTQ format were processed using Salmon (v1.9.0) for transcript quantification. Reads were pseudo-aligned to the human reference transcriptome (GRCh38, Gencode v38) and quantified at the transcript level. Transcript-level abundance estimates were imported into R and summarized to gene-level counts using the tximport package in R (v4.4.3), which aggregates transcript estimates to gene-level expression while accounting for transcript length. Genes with fewer than 10 counts across all samples were excluded from downstream analysis.

### Differential gene expression analysis

Differential gene expression analysis between postoperative PSA-event and no-event groups was performed using DESeq2 [33] in R (v4.5.2). Count data were normalized using the median-of-ratios method implemented in DESeq2. Differential-expression analyses were performed separately for tumor-adjacent benign and tumor tissues, with PCBN used for the primary discovery comparisons shown in **Figure 2A** and **Supplementary Figure 2A–B**. These PCBN discovery analyses used the original, uncorrected count data. For cross-cohort analyses, ComBat-seq was applied separately to the tumor-adjacent benign and tumor count matrices, with cohort (PCBN, BIDMC, or UM) specified as the batch variable. Because tissue-preservation method and postoperative PSA-event status were confounded with cohort, neither was included as a separate covariate. Principal component analysis was used to assess cohort integration before and after ComBat-seq correction (see **Supplementary Figure 1**). Differential expression was assessed using the Wald test with Benjamini-Hochberg false discovery rate (FDR) correction for multiple testing. Genes with adjusted p value < 0.1 and absolute log_2_ fold-change > 0.585 (equivalent to 1.5-fold change) were considered significantly differentially expressed.

### Gene set enrichment and pathway analysis

Gene Ontology (GO) and pathway enrichment analyses were performed using clusterProfiler [34]. Gene sets were obtained from the Molecular Signatures Database (MSigDB) [35], including Hallmark gene sets, GO Biological Processes, and KEGG pathways. Enrichment analysis was conducted using the enricher function with differentially expressed genes as input and all expressed genes as background. Statistical significance was determined using hypergeometric test with Benjamini-Hochberg FDR correction (adjusted p value < 0.05).

For correlation analysis between adjacent and tumor tissue, Pearson correlation coefficients were calculated for each gene using matched sample pairs. Genes with significant positive correlation (coefficient >= 0.3 and adjusted p value ≤ 0.05) were subjected to pathway enrichment analysis to identify biological processes coordinately regulated across tissue compartments. To evaluate whether correlations in matched tumor–benign pairs exceeded those expected from nonmatched samples, sample pairing was randomly permuted 100 times and the observed matched-pair correlations were compared with the resulting background distribution.

### Calculation of immune signature scores

Immune cell-specific gene sets were collected from CellMarker 2.0 [36] and CellTypist [37] reference datasets. In addition, various immune gene signatures associated with immunotherapy response prediction were obtained from Bareche et al [38]; PubMed IDs and gene members retained after expression filtering are provided in **Supplementary Table 2**. This curated panel included the B-cell gene expression signature reported by Budczies et al. [22], which was the top-ranked signature in **Figure 2B**. For each signature, genes not detected after expression filtering were omitted before scoring.

Gene signature scores were computed using the Gene Set Variation Analysis (GSVA) package (v2.0.7) [39] in R (v4.4.3) using GSVA in single-sample GSEA (ssGSEA) mode. The parameter alpha was set to 0, assigning equal weight across all ranked genes and thereby capturing the overall distribution of signature genes throughout the expression ranking rather than preferentially weighting only the highest-expressed genes. The parameter normalize = TRUE was applied to rescale enrichment scores to the range [−1, 1], enabling consistent comparisons across samples and tissue compartments. Pearson correlations between matched tumor and tumor-adjacent benign B-cell signature scores were calculated separately within the PCBN and BM cohorts.

### Cox and B-cell-associated pathway analyses

To evaluate whether immune-related signatures were associated with time to biochemical recurrence beyond established clinical risk, CAPRA-S-adjusted Cox proportional hazards models were fit separately in the PCBN and BM cohorts using the coxph() function from the survival package (version 3.8.6) in R version 4.6. Patients with an event or censoring time recorded as 0 were excluded from time-to-event analyses, yielding 108 PCBN patients with 28 biochemical-recurrence events and 118 BM patients with 45 events. Because preoperative PSA was unavailable for one of the 108 PCBN patients, the CAPRA-S-adjusted Cox models included 107 PCBN patients with 28 events. All 118 time-to-event-eligible BM patients had the clinical data required for CAPRA-S adjustment. Models were fit separately for tumor-adjacent benign and tumor tissues. Each model included a continuous immune-signature ssGSEA score and CAPRA-S score as covariates. Hazard ratios and 95% confidence intervals were estimated from CAPRA-S-adjusted Cox proportional hazards models. Wald p values were adjusted across the immune signatures tested using the Benjamini–Hochberg method.

To characterize pathways associated with adjacent benign B-cell activity, ssGSEA scores were calculated for a curated set of 38 mSigDB and other published immune, cancer-associated, and anticancer pathway signatures. Tumor-adjacent benign samples from the full cohort (n=243) were stratified into B-cell-high and B-cell-low groups using the median adjacent benign B-cell signature score. Pathway-score distributions were compared using two-tailed Wilcoxon rank-sum tests, with Benjamini–Hochberg correction across the tested pathways. Associations between the continuous adjacent benign B-cell signature score and pathway scores were evaluated using Spearman correlation, with Benjamini– Hochberg correction for multiple testing. For heatmap visualization, pathway scores were standardized across samples as row z-scores.

### Clinical risk score calculation

Two clinicopathologic risk classifications were calculated for patients with the required clinical data: a postoperative D’Amico-like [4] risk classification and the Cancer of the Prostate Risk Assessment Post-Surgical (CAPRA-S) [6] score. Because preoperative PSA was unavailable for one PCBN patient, analyses involving either classification included 122 PCBN patients and all 120 BM patients.

The postoperative D’Amico-like classification used preoperative PSA, radical-prostatectomy Gleason score [40], and pathologic T stage, applying D’Amico-derived risk thresholds:

- Low risk: PSA ≤10 ng/mL AND RP Gleason score ≤6 AND pathologic stage T1c-T2a
- Intermediate risk: PSA 10-20 ng/mL OR RP Gleason score 7 OR pathologic stage T2b
- High risk: PSA >20 ng/mL OR RP Gleason score 8-10 OR pathologic stage ≥T2c

CAPRA-S scores (range: 0-12) were calculated using six pathological and clinical variables according to published criteria:

- PSA level: 0-6 ng/mL (0 points), 6.01-10 ng/mL (1 point), 10.01-20 ng/mL (2 points), >20 ng/mL (3 points)
- Surgical margins: Negative (0 points), Positive (2 points)
- Seminal vesicle invasion: No (0 points), Yes (2 points)
- Gleason score: 2-6 (0 points), 3+4 (1 point), 4+3 (2 points), 8-10 (3 points)
- Extracapsular extension: No (0 points), Yes (1 point)
- Lymph node involvement: No (0 points), Yes (1 point)

### B cell Risk Index using Gleason score, Adjacent-tissue signature, and Diagnostic PSA for biochemical recurrencE (BRIGADE)

BRIGADE was developed using elastic net-regularized logistic regression to integrate B cell signature scores from adjacent benign tissue with clinical parameters (PSA level and Gleason score). The PCBN cohort served as the training set, and the BM cohort served as an independent validation set. Model training was performed using the glmnet engine via the caret framework in R version 4.5.2, with hyperparameters tuned through cross-validation. The optimal model used an elastic-net penalty with α = 0.90 and λ = 0.04281, reflecting predominantly Lasso-like regularization with a small L2 component. All predictors were centered and scaled prior to model fitting. The final fitted model was:

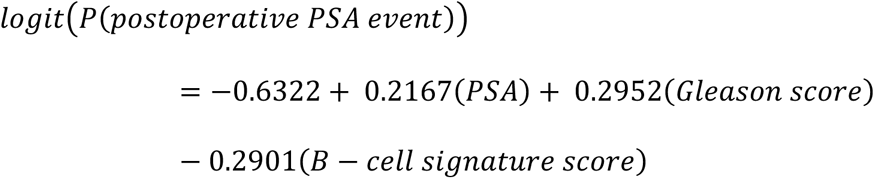

In the training cohort, model performance was evaluated using 5-fold cross-validation to provide an internal estimate of model performance. The dataset was randomly partitioned into 5 equal-sized folds, and the model was trained on 4 folds and tested on the remaining fold. This process was repeated 5 times with each fold serving as the test set exactly once. The mean AUC and AUPRC across all 5 folds were reported as the cross-validated performance estimates.

For threshold-based classification, Youden’s index [41] (sensitivity + specificity - 1) was calculated across the range of BRIGADE values in the training cohort. The threshold that maximized Youden’s index was selected as the optimal cutpoint for classifying patients into high-risk (BRIGADE > threshold) and low-risk (BRIGADE ≤ threshold) groups. Patients with complete adjacent benign B-cell signature, preoperative PSA, Gleason score, and outcome data were included in model fitting. Preoperative PSA was unavailable for one PCBN patient; therefore, BRIGADE development and cross-validation used 122 PCBN patients. For direct comparison with BRIGADE, the individual-predictor classification analyses were performed in the same 122-patient complete-case cohort. All 120 BM patients had complete data and were included in independent validation. The final PCBN model was applied to BM without coefficient refitting or threshold re-estimation.

### B-cell receptor repertoire reconstruction and Gini-index analysis

B-cell receptor repertoires were reconstructed from tumor-adjacent benign bulk RNA-sequencing data using the TRUST4 run-trust4 workflow with the hg38 BCR/TCR reference (hg38_bcrtcr.fa) and the human IMGT+C reference (human_IMGT+C.fa). For downstream B-cell receptor analyses, clonotypes annotated with immunoglobulin-chain designations beginning with IG were retained, whereas T-cell receptor clonotypes were analyzed separately and excluded from the B-cell receptor repertoire analysis. For each evaluable sample, clonal inequality was summarized using the Gini index, with higher values indicating greater concentration of the repertoire among a smaller number of clonotypes and therefore lower repertoire diversity. PCBN patients with PSA persistence or an event time recorded as 0 were excluded from the time-to-event analysis. Patients with an evaluable repertoire (n=108) were divided into Gini-high and Gini-low groups using the cohort median (0.597). Biochemical recurrence-free probability was estimated using the Kaplan–Meier method and compared using a two-sided log-rank test.

### Multiplex immunofluorescence and image analysis

For multiplex immunofluorescence verification of B-cell infiltration, formalin-fixed paraffin-embedded (FFPE) tissue sections (4 μm thick) were obtained from five BIDMC patients and five NCI patients. The multiplex panel consisted of CD19 to identify B cells, PSA to identify epithelial/tumor regions, and DAPI to identify nuclei. No-event cases were selected from the BIDMC cohort based on high adjacent-tissue B-cell signature scores in the top quartile of the corresponding RNA-seq data, whereas event-positive cases were obtained from the NCI cohort because of tissue availability. Sections were baked for 1h at 60°C. Following deparaffinization in xylenes and rehydration through graded alcohols, antigen retrieval was performed using a NxGen Decloaker (Biocare Medical) at 110°C for 15 minutes in Tris-EDTA Buffer (Abcam; ab93684), pH 9.0. Next, a thin border was drawn around the edges of each glass slide using a PAP pen. After 10-minute incubations in Background Punisher (Biocare; BP974), anti-CD19 clone EPR5906 (Abcam; ab134114) was diluted 1:250 into Renoir Red diluent (Biocare Medical; PD904) and incubated for 30mins. Slides were washed with TBST then incubated with ImmPRESS HRP Goat-anti-Rabbit IgG Polymer Reagent (Vector Laboratories; 30125) for 30mins. Slides were washed with TBST then incubated with Opal 650 (Akoya Biosciences; FP1496001KT) diluted 1:150 in 1× Plus Amplification Diluent (Akoya Biosciences; FP1498) for 1h followed by washing with TBST. Antigen retrieval was performed in citrate buffer (Biocare Medical; DV2004) pH 6.0 followed by incubation with anti-PSA clone D6B1 (Cell Signaling; 5365) for 1h. Slides were washed with TBST then incubated with Alexa Fluor 750 conjugated goat-anti-Rabbit IgG antibody (Invitrogen; A21039) for 1h followed by washing. Nuclei were stained by Spectral DAPI (Akoya; FP1490) diluted to 1× with PBS for 10mins followed by washing with TBST. Slides were wet-mounted using Prolong Gold (Invitrogen; P36980) and digitized using a Carl Zeiss AxioScan.Z1 microscope slide scanner equipped with a Plan-Apochromat 20× NA 0.8 objective. All multiplex immunofluorescence assays were performed using validated protocols on a PATH FLX autostainer (Biocare Medical).

Quantification of CD19^+^ B cell density was performed using the HALO AI platform (Indica Labs). Briefly, cell segmentation was performed using the HiPlex FL v5.2.2 algorithm based on nuclei staining in tumor-free (benign) regions or the whole tissue area. Tumor and adjacent benign tissue regions were called based on H&E staining. For nuclear segmentation, only cells between 8-57 µm^2^ were included. Next, B cells were identified based on CD19 expression 2µm beyond the nucleus while epithelial cells were identified by PSA expression 2 µm beyond nucleus. Minimum cytoplasmic intensity thresholds of 3000 and 2000 were used to define CD19 and PSA positivity, respectively, across all stained samples. B-cell and PSA-positive luminal-cell densities were calculated per unit area. Normalized B-cell density was calculated as the ratio of CD19⁺ B-cell density to PSA-positive luminal-cell density to account for differences in epithelial cellularity and tumor crowding. For each patient, B-cell nearest-neighbor distance was summarized as the mean distance from each B cell to its nearest B-cell neighbor within the analyzed benign region or whole-slide area. Analysis was performed using R version 4.2.0.

### Performance metrics: AUC and odds ratio calculation

The positive class for primary binary analyses was a postoperative PSA event. Sensitivity was defined as TP/(TP+FN), specificity as TN/(TN+FP), and the false-positive rate as FP/(FP+TN). Precision was defined as TP/(TP+FP), and recall was defined as TP/(TP+FN). AUC was calculated using the pROC package, and AUPRC was calculated using the PRROC package. AUPRC was interpreted relative to the prevalence of postoperative PSA events in each cohort.

At each selected classification threshold, the odds ratio compared the odds of a postoperative PSA event among patients classified as high risk with the odds among those classified as low risk. For descriptive purposes, the corresponding contingency-table odds ratio is (TP × TN)/(FP × FN). Odds ratios and 95% confidence intervals were estimated using logistic regression with binary high-risk/low-risk classification as the predictor. AUC values range from 0 to 1, where 1 indicates perfect discrimination, 0.5 indicates performance equivalent to random chance. AUC was calculated using the pROC package (v1.18.0) in R version 4.5.2.

Differences in normalized B-cell density and nearest-neighbor distance between event-positive and no-event cases were assessed using two-tailed Wilcoxon rank-sum tests. A normalized B-cell density cutoff was selected by maximizing Youden’s index. For exploratory time-to-event visualizations, the same 10-patient cohort was dichotomized at the cohort median for normalized B-cell density or nearest-neighbor distance; biochemical recurrence-free probability was estimated using Kaplan–Meier methods and compared using two-sided log-rank tests.

For individual predictors and composite scores including PSA, Gleason score, CAPRA-S, the postoperative D’Amico-like classification, and BRIGADE, optimal classification thresholds were determined using Youden’s index, which maximizes sensitivity + specificity − 1. The threshold corresponding to the maximum Youden’s index was used for binary classification and threshold-specific odds-ratio estimation.

### Statistical analysis and performance evaluation

Model performance was evaluated using multiple metrics. Area under the receiver operating characteristic curve (AUC) and area under the precision-recall curve (AUPRC) were calculated using the pROC [42] and PRROC [43] packages in R version 4.5.2.

Continuous variables were compared between two groups using two-tailed Wilcoxon rank-sum tests. ISUP grade-group trends were evaluated using the Cochran-Armitage test for trend. Pearson correlations were used for matched tumor-benign expression and tissue-density analyses, whereas Spearman correlations were used for associations with clinical variables and pathway scores. Kaplan-Meier curves were compared using two-sided log-rank tests. Cox-model associations were evaluated using Wald tests. Unless otherwise specified, multiple-testing correction used the Benjamini-Hochberg method.

## DISCLOSURES

### Data availability

The data underlying this article have been deposited in the Database of Genotypes and Phenotypes (https://www.ncbi.nlm.nih.gov/gap/) (phs001813.v4.p1) and the Gene Expression Omnibus (GSE183040, GSE201284, and GSE309842) (https://www.ncbi.nlm.nih.gov/geo/).

### Code availability

All original code used in this study has been deposited in GitHub at https://github.com/wbb1813/BBS_Prostate_cancer.git and is publicly available as of the date of publication. To ensure reproducibility and provide a permanent citation, the repository has also been archived in Zenodo at https://zenodo.org/records/18746612.

## Acknowledgments

The authors gratefully acknowledge the patients and the families of patients who contributed to this study.

## Contributions

Conceptualization: CCH, KDA, ABW, AGS, ER

Data curation: BW, SM, ATK

Formal Analysis: BW, SM, ATK, SK, KW

Funding acquisition: AGS, ER

Investigation: AB, SYT, RTL, NCW, ATK, KEH, SK, RL

Methodology: BW, SM, AB, ATK, RL, ABW

Project administration: AGS

Resources: OSV, CC, JS, RSM, LAK, JC, PDM, PK, LDT, AC, SG, PAP, CM, SSS, DJE, SPB

Software: BW, SM

Supervision: CT, EJF, AGS, ER

Validation: BW, SM, ATK

Visualization: BW, SM, ATK, AGS

Writing – original draft: BW, SM

Writing – review & editing: All authors

