## Supplementary for "Tumor-adjacent B cell infiltration stratifies recurrence risk in localized prostate cancer"

Binbin Wang *et al.*

- Supplementary Tables
- Supplementary Figures

**Supplementary Table 1**

| Characteristic | PCBN (N=123) | BIDMC (N=84) | UM (N=36) |
|---|---|---|---|
| Patients (N) | 123 | 84 | 36 |
| Age at RP, years, median (IQR) | 63.1 (57.4–67.7) | 64.3 (58.7–67.1) | 65.2 (59.0–68.3) |
| Pre-op PSA, ng/mL, median (IQR) | 6.7 (4.7–10.4) | 6.0 (4.5–7.9) | 7.9 (6.0–13.1) |
| **ISUP grade group, % (n)** | | | |
| Grade 1 | 6.5% (8) | 9.5% (8) | 5.6% (2) |
| Grade 2 | 33.3% (41) | 64.3% (54) | 8.3% (3) |
| Grade 3 | 40.7% (50) | 21.4% (18) | 44.4% (16) |
| Grade 4 | 4.1% (5) | 1.2% (1) | 5.6% (2) |
| Grade 5 | 15.4% (19) | 3.6% (3) | 36.1% (13) |
| **Pathologic stage, % (n)** | | | |
| pT2 | 74.0% (91) | 51.2% (43) | 36.1% (13) |
| pT3a | 15.4% (19) | 39.3% (33) | 27.8% (10) |
| pT3b | 5.7% (7) | 9.5% (8) | 36.1% (13) |
| Unknown | 4.9% (6) | 0.0% (0) | 0.0% (0) |
| **Race, % (n)** | | | |
| White | 90.2% (111) | 72.6% (61) | 91.7% (33) |
| Black | 3.3% (4) | 9.5% (8) | 5.6% (2) |
| Hispanic | 2.4% (3) | 0.0% (0) | 0.0% (0) |
| Asian | 1.6% (2) | 0.0% (0) | 2.8% (1) |
| Native American | 1.6% (2) | 0.0% (0) | 0.0% (0) |
| Hawaiian or Pacific Islander | 0.0% (0) | 1.2% (1) | 0.0% (0) |
| Other | 0.8% (1) | 0.0% (0) | 0.0% (0) |
| Unknown | 0.0% (0) | 16.7% (14) | 0.0% (0) |
| **Postoperative PSA event, % (n)** | | | |
| Event | 35.0% (43) | 13.1% (11) | 97.2% (35) |
| No event | 65.0% (80) | 86.9% (73) | 2.8% (1) |

**Supplementary Table 1. Patient characteristics by institutional cohort.** For the primary binary analyses, a postoperative PSA event included biochemical recurrence or PSA persistence. Within PCBN, 15 of the 43 event-positive cases were annotated as persistent disease in which PSA did not become undetectable after prostatectomy. Preoperative PSA was available for 122 of 123 PCBN patients; the reported PCBN PSA median and interquartile range are based on those 122 patients. **Abbreviations**: BIDMC, Beth Israel Deaconess Medical Center; IQR, interquartile range; ISUP, International Society of Urological Pathology; PCBN, Prostate Cancer Biorepository Network; PSA, prostate-specific antigen; RP, radical prostatectomy; UM, University of Michigan.

**Supplementary Table 2**

| Signature name | PMID | Gene members |
|---|---|---|
| B cell | 33520406 | BLK, CD19, FCRL2, MS4A1, TNFRSF17, TCL1A, SPIB, PNOC |
| LRRC15 CAF | 31699795 | MMP11, COL11A1, C1QTNF3, CTHRC1, COL12A1, COL10A1, COL5A2, THBS2, AEBP1, LRRC15, ITGA11 |
| Myeloid DC | 31942075 | CD1A, CD1B, CD1E, CLEC10A, CLIC2, WFDC21P |
| IRG | 28650338 | IDO1, CXCL10, CXCL9, HLA-DRA, STAT1, IFNG |
| MHC-I | 33941922 | HLA-A, HLA-B, HLA-C, B2M, TAP1, TAP2 |
| MHC-II | 33941922 | HLA-DMA, HLA-DMB, HLA-DOA, HLA-DOB, HLA-DPA1, HLA-DPB1, HLA-DQA1, HLA-DQA2, HLA-DQB1, HLA-DQB2, HLA-DRA, HLA-DRB1, HLA-DRB5 |
| Immune cells | 33033253 | ADAM21, ALDH1L2, APBB2, APOC2, ARSF, ASAH2, ASPM, BIRC5, C6orf223, CACNG1, CCL18, CD177, CD244, CDCA5, CDH1, CEACAM6, CILP2, CLDN7, CLNK, CORO7, CR2, CRTAM, CYTL1, DHRS9, DLK1, DUSP13, EIF4A2, ENTHD1, FBLN1, FBLN2, FMOD, FOLR2, FOXI1, GBP1P1, GDF1, GIMAP4, GPR31, GRIA1, GRM7, ITGA3, JMJD7, KIR2DL4, KIRREL2, KLHDC8B, KRT4, LALBA, LEF1, LINC00243, LYSMD2, MAEL, MAP2K5, MATN3, MFAP2, MKI67, MMP12, MMP9, MT1G, MUSK, MXRA8, MYL1, MYO1G, NACA2, NACA3P, NUDT10, NUPR1, OTOF, PCLAF, PKDCC, PLA2G2D, PPA2, PPP4R3C, PRPH, PRUNE2, RASL12, RIMS2, RNASE1, ROR1, RPL36AP41, RRM2, SCGB2A2, SELENOP, SH2D2A, SHC3, SLC16A3, SPATA13, SPC24, SPP1, STC1, STC2, STOML3, SYT6, TDRD15, TEAD2, TK1, TM4SF19, TMEM171, TRAF3IP2, TREM2, TRPC4, TSHZ3, TUBA8, TYMS, UBE2C, UNC80, ZNF219, ZNF462, ZNF610, ZNF880 |
| ADO | 31953314 | PPARG, CYBB, COL3A1, FOXP3, LAG3, APP, CD81, GPI, PTGS2, CASP1, FOS, MAPK1, MAPK3, CREB1 |
| CRMA | 29656892 | CRMA, MAGEA2, MAGEA2B, MAGEA3, MAGEA6, MAGEA12 |
| KDM5A TLR | 32908002 | KDM5A, RIOK1, MVB12B, HMCN1, FAM13C, PDE1A, IKZF3, ANO3 |
| APM-1 | 31767055 | PSMB5, PSMB6, PSMB7, PSMB8, PSMB9, PSMB10, TAP1, TAP2, ERAP1, ERAP2, CANX, CALR, PDIA3, TAPBP, B2M, HLA-A, HLA-B, HLA-C |
| CD8 | 30388456 | IL7R, GPR183, LMNA, NR4A3, TCF7, MGAT4A, CD55, AIM1, PER1, FOSL2, EGR1, TSPYL2, YPEL5, CSRNP1, REL, SKIL, PIK3R1, FOXP1, RGCC, PFKFB3, MYADM, ZFP36L2, USP36, TC2N, FAM177A1, BTG2, TSC22D2, FAM65B, STAT4, RGPD5, NEU1, IFRD1, PDE4B, NR4A1 |
| APM-2 | 33028693 | B2M, CALR, NLRC5, PSMB9, PSME1, PSME3, RFX5, HSP90AB1 |
| TGF | 29443960 | ACTA2, ACTG2, ADAM12, ADAM19, CNN1, COL4A1, CTGF, CTPS1, FAM101B, FSTL3, HSPB1, IGFBP3, PXDC1, SEMA7A, SH3PXD2A, TAGLN, TGFBI, TNS1, TPM1 |

| Signature name | PMID | Gene members |
|---|---|---|
| Immune Cytolytic | 25594174 | GZMA, PRF1 |
| TLS | 31942071 | CD1D, CCR6, LAT, SKAP1, CD79B, EIF1AY, RBP5, PTGDS, CETP |
| IFNG | 28650338 | CD3D, IDO1, CIITA, CD3E, CCL5, GZMK, CD2, HLA-DRA, CXCL13, IL2RG, NKG7, HLA-E, CXCR6, LAG3, TAGAP, CXCL10, STAT1, GZMB |

**Supplementary Table 2. Immune-related signatures evaluated in tumor-adjacent benign transcriptomes.** Signature labels correspond to **Figure 2B**. PubMed IDs and gene members retained after expression filtering and used for ssGSEA scoring are listed.

**Supplementary Table 3**

| Pathway signature | B-cell high, median ssGSEA score | B-cell low, median ssGSEA score | Difference, high - low | p value | FDR-adjusted p value | Higher median group |
|---|---|---|---|---|---|---|
| CCP Prolaris Cuzick2011 | 0.284 | −0.275 | 0.559 | $7.66 \times 10^{-17}$ | $2.91 \times 10^{-15}$ | B-cell high |
| HALLMARK COMPLEMENT | 0.313 | −0.159 | 0.472 | $3.88 \times 10^{-16}$ | $6.15 \times 10^{-15}$ | B-cell high |
| RB Loss Chen2019 | 0.349 | −0.206 | 0.556 | $4.85 \times 10^{-16}$ | $6.15 \times 10^{-15}$ | B-cell high |
| HALLMARK IL6 JAK STAT3 SIGNALING | 0.296 | −0.245 | 0.541 | $9.61 \times 10^{-16}$ | $9.13 \times 10^{-15}$ | B-cell high |
| HRD Signature | 0.396 | −0.009 | 0.405 | $1.54 \times 10^{-15}$ | $1.17 \times 10^{-14}$ | B-cell high |
| HALLMARK KRAS SIGNALING UP | 0.276 | −0.143 | 0.419 | $3.29 \times 10^{-15}$ | $2.09 \times 10^{-14}$ | B-cell high |
| HALLMARK INFLAMMATORY RESPONSE | 0.300 | −0.230 | 0.530 | $8.40 \times 10^{-15}$ | $4.56 \times 10^{-14}$ | B-cell high |
| HALLMARK INTERFERON GAMMA RESPONSE | 0.291 | −0.233 | 0.524 | $1.33 \times 10^{-14}$ | $6.30 \times 10^{-14}$ | B-cell high |
| Ki67 Proliferation | 0.253 | −0.262 | 0.514 | $1.47 \times 10^{-13}$ | $6.22 \times 10^{-13}$ | B-cell high |
| HALLMARK ANGIOGENESIS | 0.285 | −0.165 | 0.450 | $2.44 \times 10^{-13}$ | $9.26 \times 10^{-13}$ | B-cell high |
| HALLMARK EPITHELIAL MESENCHYMAL TRANSITION | 0.315 | −0.131 | 0.447 | $1.45 \times 10^{-12}$ | $5.00 \times 10^{-12}$ | B-cell high |
| HALLMARK G2M CHECKPOINT | 0.279 | −0.237 | 0.516 | $2.92 \times 10^{-12}$ | $9.26 \times 10^{-12}$ | B-cell high |
| HALLMARK E2F TARGETS | 0.297 | −0.132 | 0.429 | $5.14 \times 10^{-12}$ | $1.50 \times 10^{-11}$ | B-cell high |
| HALLMARK MITOTIC SPINDLE | 0.312 | −0.151 | 0.463 | $1.52 \times 10^{-11}$ | $4.14 \times 10^{-11}$ | B-cell high |
| Decipher GC Erho2013 | 0.236 | −0.162 | 0.398 | $3.50 \times 10^{-11}$ | $8.86 \times 10^{-11}$ | B-cell high |
| HALLMARK INTERFERON ALPHA RESPONSE | 0.282 | −0.207 | 0.490 | $9.93 \times 10^{-11}$ | $2.36 \times 10^{-10}$ | B-cell high |
| HALLMARK APOPTOSIS | 0.286 | −0.145 | 0.431 | $1.39 \times 10^{-10}$ | $3.10 \times 10^{-10}$ | B-cell high |
| HALLMARK TNFA SIGNALING VIA NFKB | 0.318 | −0.241 | 0.559 | $2.08 \times 10^{-10}$ | $4.39 \times 10^{-10}$ | B-cell high |
| ERG Fusion Signature | 0.272 | −0.116 | 0.388 | $2.39 \times 10^{-10}$ | $4.79 \times 10^{-10}$ | B-cell high |
| HALLMARK HYPOXIA | 0.266 | −0.121 | 0.387 | $6.97 \times 10^{-10}$ | $1.32 \times 10^{-9}$ | B-cell high |
| HALLMARK HEDGEHOG SIGNALING | 0.201 | −0.139 | 0.340 | $1.32 \times 10^{-7}$ | $2.39 \times 10^{-7}$ | B-cell high |
| HALLMARK P53 PATHWAY | 0.276 | −0.117 | 0.393 | $3.81 \times 10^{-7}$ | $6.57 \times 10^{-7}$ | B-cell high |
| HALLMARK NOTCH SIGNALING | 0.292 | −0.205 | 0.497 | $3.65 \times 10^{-6}$ | $6.03 \times 10^{-6}$ | B-cell high |

| Pathway signature | B-cell high, median ssGSEA score | B-cell low, median ssGSEA score | Difference, high - low | p value | FDR-adjusted p value | Higher median group |
|---|---|---|---|---|---|---|
| HALLMARK WNT BETA CATENIN SIGNALING | 0.266 | −0.129 | 0.395 | $1.79 \times 10^{-5}$ | $2.79 \times 10^{-5}$ | B-cell high |
| HALLMARK GLYCOLYSIS | 0.236 | −0.116 | 0.352 | $1.84 \times 10^{-5}$ | $2.79 \times 10^{-5}$ | B-cell high |
| HALLMARK FATTY ACID METABOLISM | 0.284 | 0.020 | 0.264 | $2.93 \times 10^{-5}$ | $4.29 \times 10^{-5}$ | B-cell high |
| PTEN Loss Proxy | 0.329 | −0.030 | 0.358 | $1.28 \times 10^{-4}$ | $1.80 \times 10^{-4}$ | B-cell high |
| MMR Signature | 0.274 | −0.025 | 0.299 | $1.45 \times 10^{-4}$ | $1.97 \times 10^{-4}$ | B-cell high |
| HALLMARK MTORC1 SIGNALING | 0.300 | −0.031 | 0.331 | $4.03 \times 10^{-4}$ | $5.28 \times 10^{-4}$ | B-cell high |
| HALLMARK MYC TARGETS V1 | 0.317 | −0.226 | 0.544 | $4.56 \times 10^{-4}$ | $5.77 \times 10^{-4}$ | B-cell high |
| HALLMARK MYC TARGETS V2 | 0.207 | −0.139 | 0.345 | $5.32 \times 10^{-4}$ | $6.53 \times 10^{-4}$ | B-cell high |
| HALLMARK DNA REPAIR | 0.284 | −0.092 | 0.376 | 0.00121 | 0.0014 | B-cell high |
| NE Score Beltran2016 | 0.106 | −0.095 | 0.201 | 0.00121 | 0.0014 | B-cell high |
| HALLMARK OXIDATIVE PHOSPHORYLATION | 0.347 | −0.029 | 0.376 | 0.00992 | 0.0111 | B-cell high |
| HALLMARK ANDROGEN RESPONSE | 0.179 | 0.005 | 0.174 | 0.0114 | 0.0123 | B-cell high |
| PSMA FOLH1 Module | −0.067 | 0.068 | −0.135 | 0.131 | 0.138 | B-cell low |
| AR Score Beltran2016 | 0.207 | 0.208 | −0.0004 | 0.348 | 0.357 | B-cell low |
| HALLMARK CHOLESTEROL HOMEOSTASIS | 0.112 | 0.014 | 0.098 | 0.408 | 0.408 | B-cell high |

**Supplementary Table 3. Comparison of pathway activity between tumor-adjacent benign B-cell-high and B-cell-low tissues.** Samples from the full cohort (n=243) were divided at the median adjacent benign B-cell signature score. Pathway ssGSEA scores were compared using two-tailed Wilcoxon rank-sum tests. Difference denotes median (B-cell high) minus median (B-cell low); the higher-median group is reported irrespective of statistical significance. FDR-adjusted p values are shown for multiple pathway comparisons. **Abbreviations:** FDR, false discovery rate; ssGSEA, single-sample gene set enrichment analysis.

**Supplementary Table 4**

| Pathway signature | Spearman rho | p value | FDR-adjusted p value | Association with B-cell score |
|---|---|---|---|---|
| CCP Prolaris Cuzick2011 | 0.607 | $6.10 \times 10^{-26}$ | $2.32 \times 10^{-24}$ | Positive |
| HALLMARK IL6 JAK STAT3 SIGNALING | 0.533 | $2.62 \times 10^{-19}$ | $4.92 \times 10^{-18}$ | Positive |
| RB Loss Chen2019 | 0.531 | $3.88 \times 10^{-19}$ | $4.92 \times 10^{-18}$ | Positive |
| HALLMARK INFLAMMATORY RESPONSE | 0.527 | $7.75 \times 10^{-19}$ | $7.36 \times 10^{-18}$ | Positive |
| HALLMARK INTERFERON GAMMA RESPONSE | 0.497 | $1.29 \times 10^{-16}$ | $9.79 \times 10^{-16}$ | Positive |
| HALLMARK COMPLEMENT | 0.481 | $1.46 \times 10^{-15}$ | $9.22 \times 10^{-15}$ | Positive |
| HALLMARK KRAS SIGNALING UP | 0.464 | $2.07 \times 10^{-14}$ | $1.12 \times 10^{-13}$ | Positive |
| Ki67 Proliferation | 0.460 | $3.38 \times 10^{-14}$ | $1.61 \times 10^{-13}$ | Positive |
| HRD Signature | 0.459 | $4.15 \times 10^{-14}$ | $1.75 \times 10^{-13}$ | Positive |
| HALLMARK ANGIOGENESIS | 0.414 | $1.52 \times 10^{-11}$ | $5.76 \times 10^{-11}$ | Positive |
| HALLMARK TNFA SIGNALING VIA NFKB | 0.400 | $8.58 \times 10^{-11}$ | $2.96 \times 10^{-10}$ | Positive |
| HALLMARK EPITHELIAL MESENCHYMAL TRANSITION | 0.397 | $1.23 \times 10^{-10}$ | $3.90 \times 10^{-10}$ | Positive |
| Decipher GC Erho2013 | 0.387 | $3.87 \times 10^{-10}$ | $1.13 \times 10^{-9}$ | Positive |
| HALLMARK INTERFERON ALPHA RESPONSE | 0.384 | $5.34 \times 10^{-10}$ | $1.45 \times 10^{-9}$ | Positive |
| HALLMARK G2M CHECKPOINT | 0.378 | $9.93 \times 10^{-10}$ | $2.52 \times 10^{-9}$ | Positive |
| HALLMARK E2F TARGETS | 0.357 | $9.16 \times 10^{-9}$ | $2.18 \times 10^{-8}$ | Positive |
| HALLMARK MITOTIC SPINDLE | 0.340 | $5.04 \times 10^{-8}$ | $1.13 \times 10^{-7}$ | Positive |
| ERG Fusion Signature | 0.336 | $7.81 \times 10^{-8}$ | $1.65 \times 10^{-7}$ | Positive |
| HALLMARK APOPTOSIS | 0.319 | $3.69 \times 10^{-7}$ | $7.38 \times 10^{-7}$ | Positive |
| HALLMARK HYPOXIA | 0.285 | $6.01 \times 10^{-6}$ | $1.14 \times 10^{-5}$ | Positive |
| HALLMARK HEDGEHOG SIGNALING | 0.247 | $9.45 \times 10^{-5}$ | $1.71 \times 10^{-4}$ | Positive |
| HALLMARK P53 PATHWAY | 0.210 | $9.45 \times 10^{-4}$ | 0.00163 | Positive |
| HALLMARK NOTCH SIGNALING | 0.208 | 0.00109 | 0.00181 | Positive |
| NE Score Beltran2016 | 0.177 | 0.00564 | 0.00872 | Positive |
| HALLMARK WNT BETA CATENIN SIGNALING | 0.176 | 0.00574 | 0.00872 | Positive |
| HALLMARK CHOLESTEROL HOMEOSTASIS | −0.167 | 0.0091 | 0.0133 | Negative |
| PSMA FOLH1 Module | −0.144 | 0.0241 | 0.034 | Negative |
| MMR Signature | 0.122 | 0.0576 | 0.0782 | Positive |
| PTEN Loss Proxy | 0.120 | 0.0616 | 0.0807 | Positive |
| HALLMARK GLYCOLYSIS | 0.112 | 0.0821 | 0.104 | Positive |

| Pathway signature | Spearman rho | p value | FDR-adjusted p value | Association with B-cell score |
|---|---|---|---|---|
| HALLMARK MYC TARGETS V2 | 0.110 | 0.0863 | 0.106 | Positive |
| AR Score Beltran2016 | −0.094 | 0.145 | 0.172 | Negative |
| HALLMARK MYC TARGETS V1 | 0.066 | 0.304 | 0.348 | Positive |
| HALLMARK FATTY ACID METABOLISM | 0.065 | 0.311 | 0.348 | Positive |
| HALLMARK MTORC1 SIGNALING | 0.057 | 0.379 | 0.412 | Positive |
| HALLMARK OXIDATIVE PHOSPHORYLATION | −0.041 | 0.52 | 0.549 | Negative |
| HALLMARK ANDROGEN RESPONSE | −0.036 | 0.577 | 0.593 | Negative |
| HALLMARK DNA REPAIR | 0.028 | 0.665 | 0.665 | Positive |

**Supplementary Table 4. Correlations between continuous adjacent benign B-cell signature scores and pathway activity.** Spearman correlations were calculated between the continuous adjacent benign B-cell ssGSEA score and pathway ssGSEA scores in tumor-adjacent benign tissues from the full cohort (n=243). Positive rho values indicate increasing pathway activity with increasing B-cell signature score, whereas negative rho values indicate an inverse association. FDR-adjusted p values are shown for multiple pathway comparisons. **Abbreviations:** FDR, false discovery rate; ssGSEA, single-sample gene set enrichment analysis.

### Supplementary Figure 1

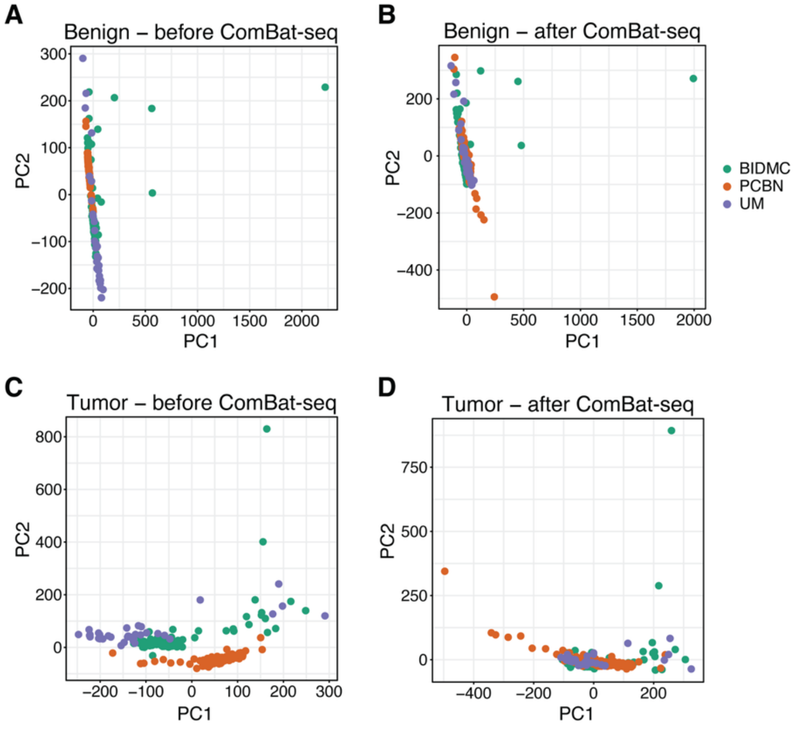


**Supplementary Figure 1. Batch correction of transcriptomic data across cohorts. A–B.** Principal component analysis of tumor-adjacent benign transcriptomic profiles before (**A**) and after (**B**) batch correction using ComBat-seq, with points colored by cohort (PCBN, BIDMC, and UM). **C–D.** Principal component analysis of tumor transcriptomic profiles before (**C**) and after (**D**) batch correction using ComBat-seq. Each point represents one sample (tumor-adjacent benign, n=243; tumor, n=243).

### Supplementary Figure 2

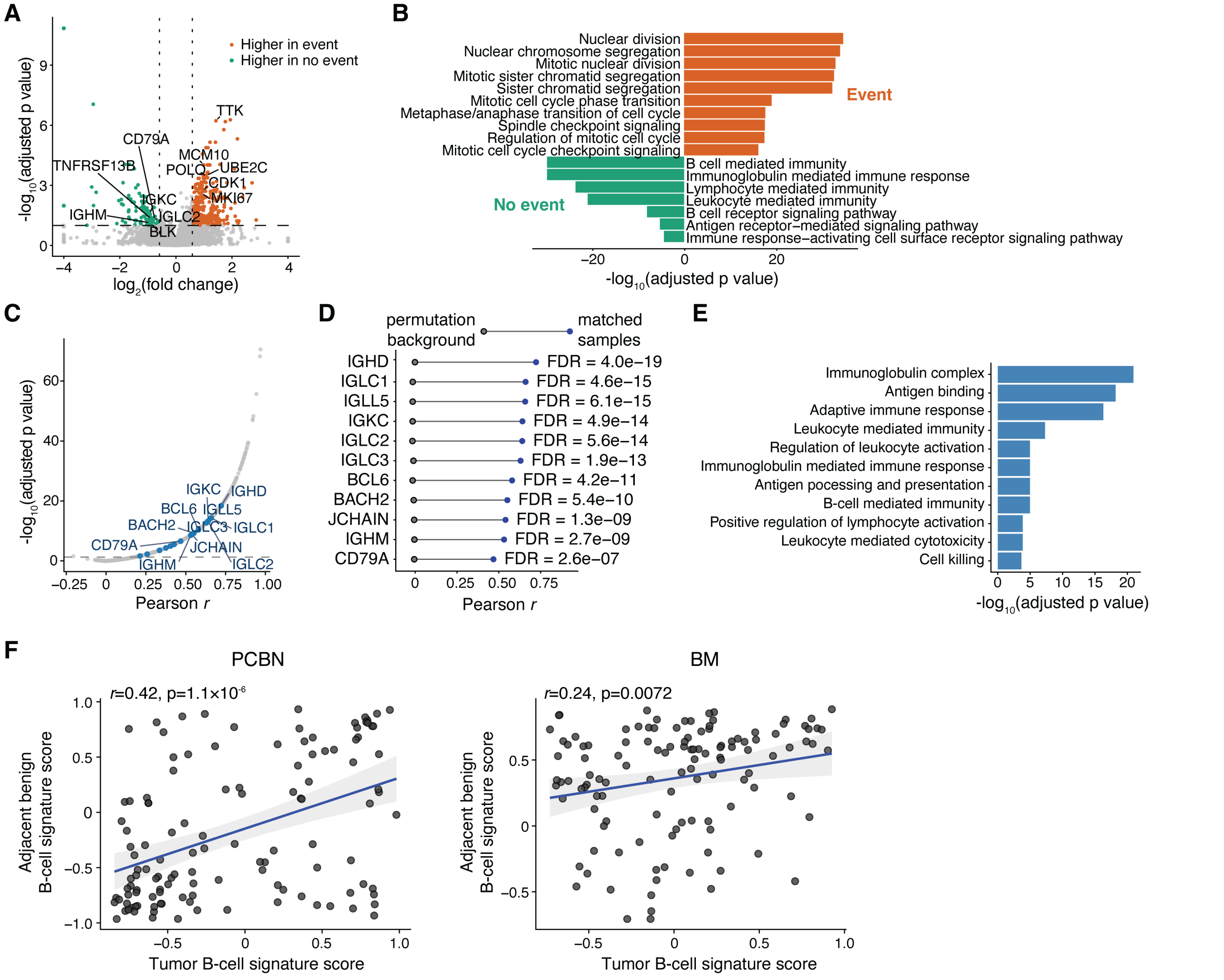


**Supplementary Figure 2. Tumor transcriptional programs and correlations between matched tumor and tumor-adjacent benign tissues. A.** Volcano plot of differential gene expression comparing tumor tissues from patients with and without a postoperative PSA event in the PCBN cohort (n=123). Genes with higher expression in postoperative PSA-event cases are shown in orange, and genes with higher expression in no-event cases are shown in green. Horizontal and vertical dashed lines indicate the adjusted p-value threshold of 0.1 and absolute $\log_2$ fold-change threshold of 0.585, respectively. Representative B-cell-related and cell-cycle-related genes are labeled. **B.** Pathway enrichment analysis of genes with higher expression in postoperative PSA-event cases (orange) or no-event cases (green). Bar length represents $-\log_{10}$(adjusted p value), with bars extending in opposite directions according to postoperative PSA-event status. **C.** Pearson correlation coefficients for gene expression between matched tumor and tumor-adjacent benign tissues. Selected B-cell-related genes are labeled in blue; the horizontal dashed line indicates the adjusted p-value threshold of 0.05. **D.** Pearson correlations for selected B-cell-related genes in matched tumor–benign sample pairs compared with background correlations obtained by random permutation of sample pairing. Blue points indicate matched samples, gray points indicate permutation background, and FDR-adjusted p values are shown for each gene. **E.** Pathway enrichment analysis of genes positively correlated between matched tumor and tumor-adjacent benign tissues. Bar length represents $-\log_{10}$(adjusted p value). **F.** Pearson correlations between B-cell signature scores in matched tumor and tumor-adjacent benign tissues from the PCBN (left; n=123) and BM (right; n=120) cohorts. Solid lines indicate linear fits, and shaded regions indicate 95% confidence intervals. Pearson correlation coefficients and two-sided p values are shown.

**Supplementary Figure 3**

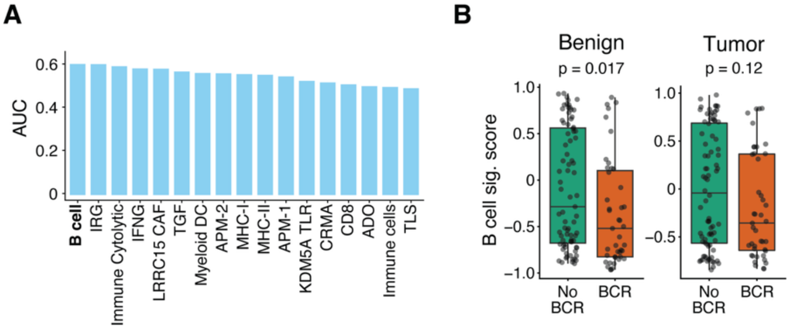


**Supplementary Figure 3. Sensitivity analysis of the PCBN cohort after excluding patients with PSA persistence.** Patients with PSA persistence or recurrence time recorded as 0 were excluded, resulting in a sensitivity cohort of 108 patients (80 without subsequent biochemical recurrence and 28 with biochemical recurrence). **A.** Area under the receiver operating characteristic curve (AUC) for a curated set of immune-related gene signatures applied to tumor-adjacent benign tissues. PubMed IDs and gene members are provided in **Supplementary Table 2**. **B.** B-cell signature enrichment scores in tumor-adjacent benign and tumor tissues, stratified by subsequent biochemical-recurrence status. P values were determined using two-tailed Wilcoxon rank-sum tests. Box plots indicate the median, interquartile range, and whiskers extending to 1.5× the interquartile range. BCR: biochemical recurrence.

### Supplementary Figure 4

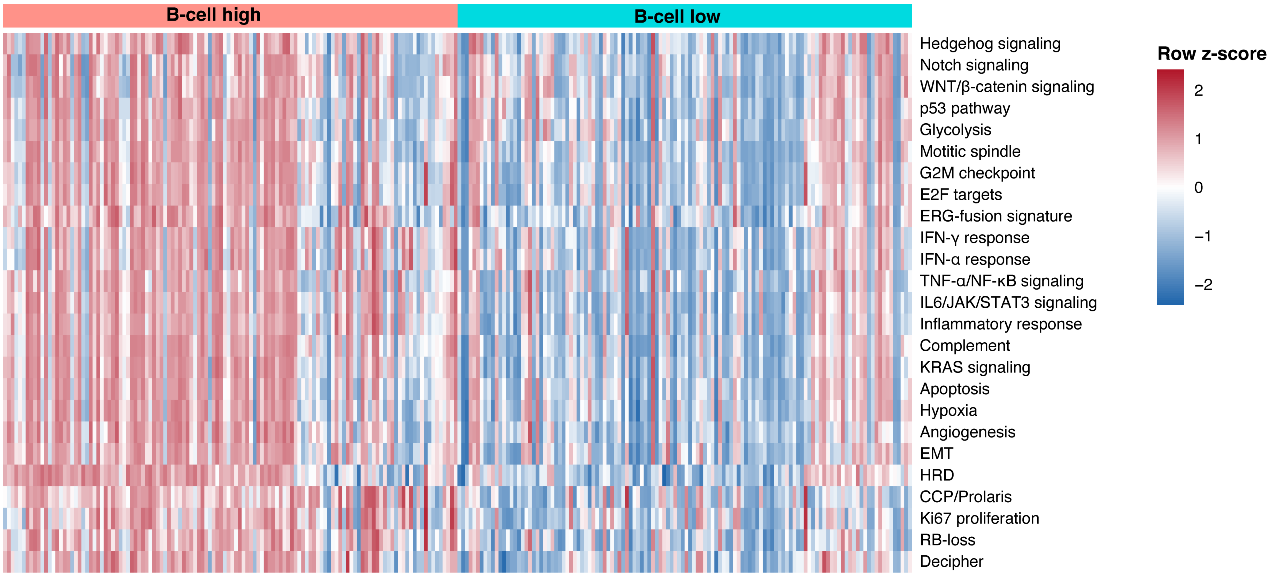


**Supplementary Figure 4. Pathway programs associated with adjacent benign B-cell activity.** Heatmap of pathway ssGSEA scores in tumor-adjacent benign tissues from the full cohort (n=243). Samples were stratified into B-cell-high and B-cell-low groups using the median adjacent benign B-cell signature score. Rows represent curated immune, cancer-associated, and anti-cancer pathway signatures, and columns represent individual samples. Values are displayed as row z-scores. Pathways are shown in the order obtained by hierarchical clustering. Complete group-comparison and continuous-correlation results are provided in **Supplementary Tables 3 and 4**.

### Supplementary Figure 5

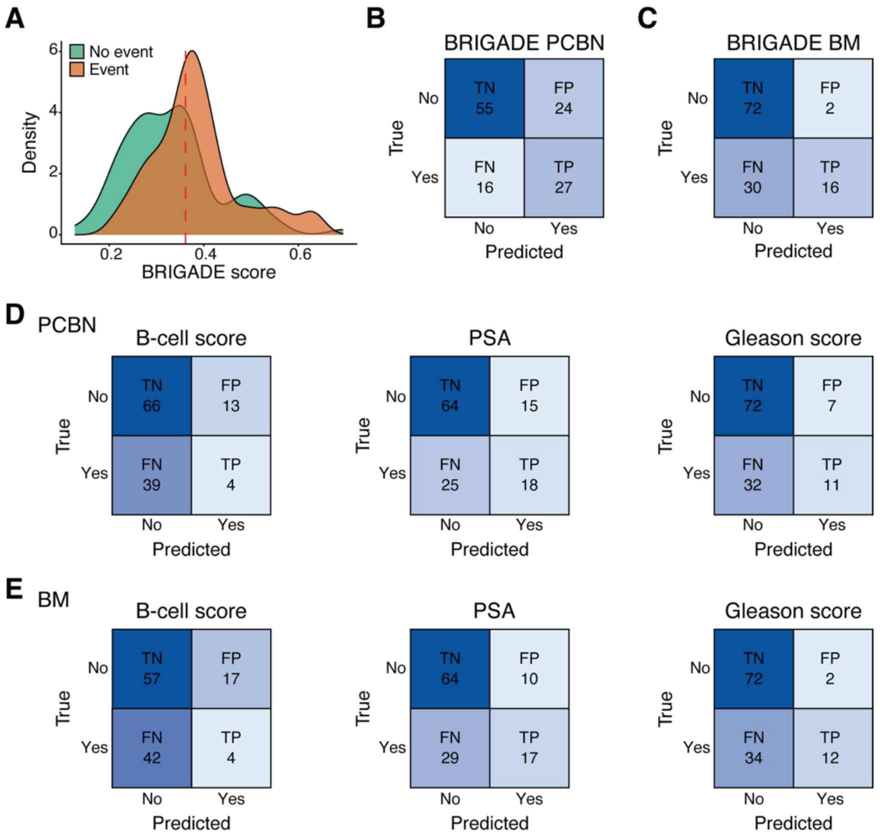


**Supplementary Figure 5. Classification performance of BRIGADE and individual predictors. A.** Density distributions of BRIGADE scores by postoperative PSA-event status in the PCBN complete-case cohort (n=122). **B–C.** Confusion matrices for BRIGADE-based classification using the fixed cutoff of 0.361 in PCBN (**B**; n=122) and BM (**C**; n=120). **D–E.** Confusion matrices for classifications based on the adjacent benign B-cell signature score, preoperative PSA, and Gleason score in the same PCBN complete-case cohort (**D**; n=122) and BM (**E**; n=120). "Yes" indicates a postoperative PSA event. TN: true negative; FP: false positive; FN: false negative; TP: true positive.

**Supplementary Figure 6**

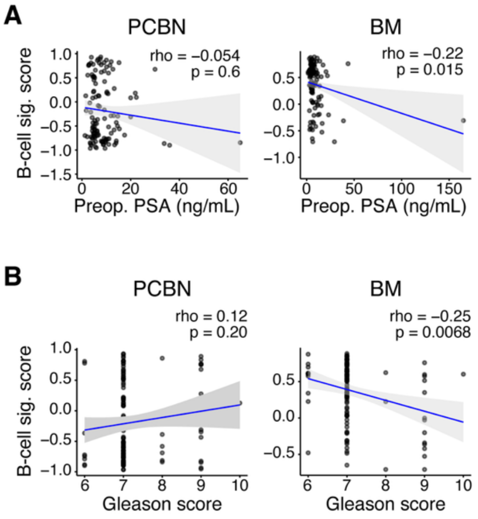


**Supplementary Figure 6. Correlations between the adjacent benign B-cell signature score and clinical variables. A.** Spearman correlations between the adjacent benign B-cell signature score and preoperative PSA in PCBN (n=122) and BM (n=120). **B.** Spearman correlations between the adjacent benign B-cell signature score and radical-prostatectomy Gleason score in PCBN (n=123) and BM (n=120). Each point represents one patient. Solid lines and shaded regions indicate linear fits and 95% confidence intervals for visualization. Spearman correlation coefficients (rho) and two-sided p values are shown in each panel.

### Supplementary Figure 7

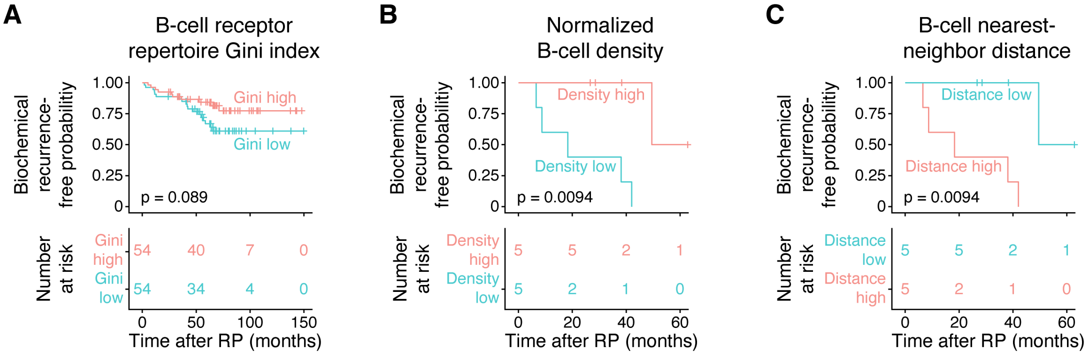


**Supplementary Figure 7. Exploratory time-to-event analyses of B-cell receptor repertoire clonality and tissue-level B-cell features. A.** Kaplan–Meier analysis of biochemical recurrence-free probability in PCBN patients with an evaluable B-cell receptor repertoire Gini index after excluding patients with PSA persistence or recurrence time recorded as 0 (n=108). Patients were stratified into Gini-high and Gini-low groups using the cohort median. **B–C.** Kaplan–Meier analyses of the same 10-patient tissue-verification cohort shown in **Figure 5C–D**, stratified using the cohort median for normalized B-cell density (**B**) or B-cell nearest-neighbor distance (**C**). Censoring is indicated by tick marks, and numbers at risk are shown below each plot. P values were determined using two-sided log-rank tests. RP: radical prostatectomy.
